# Pooled Surveillance Testing Program for Asymptomatic SARS-CoV-2 Infections in K-12 Schools and Universities

**DOI:** 10.1101/2021.02.09.21251464

**Authors:** Chongfeng Bi, Rachelle Mendoza, Hui-Ting Cheng, Gil Pagapas, Elmer Gabutan, Nadia Khan, Helen Hoxie, Kelly Holmes, Nicholas Gao, Raychel Lewis, Huaien Wang, Daniel Neumann, Angela Chan, Meril Takizawa, James Lowe, Xiao Chen, Brianna Kelly, Aneeza Asif, Keena Barnes, Nusrat Khan, Brandon May, Mecca Wright, Tasnim Chowdhury, Gabriella Pollonini, Nourelhoda Gouda, Chante Guy, Candice Gordon, Nana Ayoluwa, Elvin Colon, Noah Miller-Medzon, Shanique Jones, Rauful Hossain, Arabia Dodson, Meimei Wang, Miranda McGaskey, Ana Vasileva, Sean Seibel, James Connolly, Michele Esposito, Jane Kim, Andrew E. Lincoln, Robby Sikka, Anne L. Wyllie, Ethan M. Berke, Jenny Libien, Matthew Pincus, Prem K. Premsrirut

## Abstract

The negative impact of continued school closures during the height of the COVID-19 pandemic warrants the establishment of new cost-effective strategies for surveillance and screening to safely reopen and monitor for potential in-school transmission. Here, we present a novel approach to increase the availability of repetitive and routine Covid-19 testing that may ultimately reduce the overall viral burden in the community. We describe implementation of a testing program that included students, faculty and staff from K-12 schools and universities participating in the SalivaClear™ pooled surveillance method (Mirimus Clinical Labs, Brooklyn, NY). Over 400,000 saliva specimens were self-collected from students, faculty and staff from 93 K-12 schools and 18 universities and tested in pools of up to 24 samples over a 20-week period during this pandemic. Peaks of positive cases were seen in the days following the Halloween, Thanksgiving and New Year holidays. Pooled testing did not significantly alter the sensitivity of the molecular assay in terms of both qualitative (100% detection rate on both pooled and individual samples) and quantitative (comparable cycle threshold (CT) values between pooled and individual samples) measures. Pooling samples substantially reduced the costs associated with PCR testing and allowed schools to rapidly assess transmission and adjust prevention protocols as necessary. By establishing low-cost, weekly testing of students and faculty, pooled saliva analysis enabled schools to determine whether transmission had occurred, make data-driven decisions, and adjust safety protocols. Pooled testing is a fundamental component to the reopening of schools, minimizing transmission among students and faculty.

## Introduction

The number of daily tests for SARS-CoV-2 in the United States increased from approximately 400,000 in the summer of 2020, to 1 million daily tests in September, to 2 million in January 2021^1^. Despite this increase, access to testing remains limited. New strategies for surveillance and screening are needed to increase the availability of testing and to reduce the overall viral burden in the community while monitoring for virus resurgence. Since asymptomatic and pre-symptomatic spread are believed to be the main drivers of SARS-CoV-2 spread^2^, surveillance testing of asymptomatic individuals to screen for the presence of SARS-CoV-2 has been advocated by public health officials for interrupting chains of transmission. Despite the advent of Covid-19 vaccines, continued testing is required to monitor the spread of new SARS-CoV-2 variants and for assessing the efficacy of current vaccination strategies^3^. While RT-PCR remains the gold standard method for detecting SARS-CoV-2, cost-effective strategies are desperately needed to permit mass testing efforts.

Pooling samples for surveillance testing for SARS-CoV-2 is one effective approach for minimizing the resources required and providing a “multiplier” for existing testing frameworks^4-8^. This approach for testing individuals is also not a new idea. Pooled testing methods were integral during HIV epidemics^9^ to reduce the costs of testing. In addition, pooled testing of blood donations has been routinely used for hepatitis B virus, hepatitis C virus and HIV screening in blood banks^10^. While many pooled testing strategies have been proposed to combat the limited testing capacity during the recent pandemic, few have been successfully implemented into testing labs – and none on a large scale. Importantly, those that have, have primarily focused on swabs, which have been shown to miss nearly 15% of cases when self-collected and or miss samples with low viral loads^11,12^. Saliva in comparison, can be reliably self-collected, reducing the burden on healthcare workers and provides a sample in a format that can readily be pooled^6-8^.

Here, we describe the real-world implementation of pooled testing in K-12 schools and universities to provide an affordable approach to surveillance testing of entire school populations.

## Methods

### Study Subjects

Samples were obtained from K-12 and university students, faculty and staff members attending educational institutions participating in the SalivaClear™ (Mirimus Clinical Labs, Brooklyn, NY) pooled SARS-CoV-2 surveillance from August 27, 2020 to January 10, 2021. SalivaClear™ is a pooled testing program that utilizes a laboratory-developed saliva-based, qualitative, real time, reverse transcription–polymerase chain reaction (RT-PCR) test surveillance model adopted by numerous school districts in four northeast states. Written informed consent for Covid-19 testing was obtained from each participant from all institutions prior to the start of the surveillance program. The SUNY Downstate Health Sciences University Institutional Review Board (IRB) reviewed and approved the study protocol (IRB #1232938-3).

### Specimen Collection and Processing

Students, faculty and staff were tested 1-2 times per week using a twenty-four to one (24:1) SalivaClear™ pooled testing program. The specimen collection was performed under supervision of a trained representative from each organization. The training of the specimen collection supervisor was provided by laboratory professionals and involved a video tutorial followed by a live demonstration and FAQs session. Subjects were asked to collect saliva by passively drooling through a saliva collection device inserted into a 2-ml dry, sterile tube, each with two barcodes for accurate specimen identification. The saliva specimens were grouped by organizations’ “common exposure environment” guidelines in sets of up to 24 individuals sharing the same exposure environment (i.e., same classroom, same office). The specimens were packed and shipped according to current CDC guidelines on packing and shipping of infectious specimens. The median shipment time was 12 hours (range: 8 to 24 hours).

### Pooling of Saliva

Each set of 24 specimens was scanned into the accessioning system that creates a permanent snapshot of the location of individual tubes within that set. A designated “pool tube” was scanned with the 24 tubes. The individual tubes were automatically decapped and placed on an automated liquid handler. To decrease the viscosity of the saliva specimens, 50 µl of 0.4M 1,4-dithiotheirtol (DTT) was added and mixed with the saliva to facilitate pipetting. Following mixing, 200 µl of saliva from each individual specimen was pipetted into the designated 4-ml pre-barcoded “pool tube.”

### RNA Extraction and SARS-CoV-2 Assay

RNA extraction on pooled saliva specimens was conducted using the MagMAX Viral/Pathogen Nucleic acid isolation kit automated on the KingFisher Flex Purification system (Thermo Fisher Scientific, Waltham, MA) following manufacturer’s protocol. The RT-PCR including cDNA synthesis and PCR amplification of the target sequences was performed in triplicate on the QuantStudio 7 Pro Real-Time Flex PCR system (Applied Biosystems, Foster City, CA). The assay utilized the primer and probe sets included in the TaqPath COVID-19 Combo Kit (FDA EUA on March 13, 2020) (Thermo Fisher Scientific, Waltham, MA). These primer and probe sets were designed to amplify and detect three regions of the SARS-CoV-2 single stranded RNA genome: the Orf1ab, N and S genes. The data was analyzed by Applied Biosystems Design and Analysis Software version 2.3.4, which was used only for guidance on pooled specimens. The limit of detection (LOD) of the RT-PCR assay was validated using whole heat-inactivated SARS-CoV-2 (ATCC® VR-1986HK™, Manassas, VA) virus spiked into known negative saliva. Detection of 2 or more gene targets with cycle threshold (CT) values of at most 35 indicated a positive individual specimen, whereas detection of only 1 of the three viral gene targets was deemed inconclusive. For pooled specimens, amplification of a single target was deemed positive and resulted in reflex testing for deconvolution to identify the positive specimen(s) within that pool. The LOD for a pooled specimen was determined by the viral load that triggered reflexing in 20/20 replicates. Results were entered into an electronic health records portal and reported to the individual schools and state departments of health.

### Statistical Methods

Statistical analyses were performed in RStudio v1.3.959 (http://www.rstudio.com/), using R v4.0.2 (https://www.r-project.org/) and data visualization was performed by R package ggplot2 and ggpubr (https://rpkgs.datanovia.com/ggpubr)^13,14^. Student’s t test was used to determine significant differences in the number of positive pools and individuals at different time periods. A two-sided p value of less than 0.05 (2-tailed) was considered statistically significance. All values are shown without correction for multiple testing. The widths of the confidence intervals have not been adjusted for multiple comparisons; therefore, intervals are not used to infer definite associations. The raw data set for this study is available on a secure server hosted by Mirimus Clinical Labs, Brooklyn, NY.

## Results

Our goal was to develop a cost-efficient screening and surveillance Covid-19 testing strategy to be utilized in settings where frequent testing among asymptomatic individuals may be required to prevent early transmission. We first determined the limit of detection (LOD) of the RT-PCR assay using validated whole heat-inactivated SARS-CoV-2 virus spiked into known negative saliva (Table 1). Our results showed consistent detection of all three viral gene targets at the lowest concentration of 1 virus RNA copy/µl of sample. This was confirmed in 20 replicates of the individual sample (100 µl sample spiked with 1 and 3 virus RNA copy/µl) (Table 1). The same LOD was analyzed in a 1:24 pooled sample, where 200 µl of a sample spiked with 1-12 virus RNA copy/µl of sample was mixed with 23 x 200 µl (4600 µl total volume) of RT PCR negative saliva matrix (Supplementary Table 1). Although there was not consistent detection of the three viral targets in the 1 virus RNA copy/µl pooled specimen, in 20 out of 20 replicates a single gene amplification was seen which triggered reflex 100% of the time (Table 2; Supplementary Table 1). We repeated the LOD assay using a single sample with 6 virus RNA copy/µl of sample combined with 23 negative saliva samples and demonstrate that the pooled specimen was positive in 20/20 cases, with at least 2 of 3 genes amplified in all replicates (Table 2). We therefore set our LOD for a pooled specimen of 24 at 6 virus RNA copy/µl of sample.

**Table 1.** Limit of detection (LOD) finding study using known concentrations of ATCC VR-1986HK whole inactivated virus spiked into negative saliva. The stock concentration provided by ATCC was 4.5×10^5^ GCE/µL.

| Viral RNA<br>Copy/μl | N | Mean Ct values (SD) | | | | Positivity |
| --- | --- | --- | --- | --- | --- | --- |
|  |  | MS2 Phage | N gene | ORF1ab | S gene |  |
| 100 | 3 | 26.99 (0.31) | 25.45 (0.51) | 25.97 (0.30) | 25.10 (0.71) | 100% |
| 50 | 3 | 27.19 (0.35) | 26.62 (0.55) | 27.45 (0.79) | 27.05 (0.62) | 100% |
| 25 | 3 | 26.89 (0.47) | 26.40 (0.51) | 27.31 (0.55) | 26.84 (0.50) | 100% |
| 12 | 3 | 27.13 (0.33) | 27.22 (0.40) | 28.11 (0.41) | 27.56 (0.33) | 100% |
| 6 | 7 | 26.05 (0.65) | 27.81 (0.46) | 30.29 (1.46) | 28.07 (0.66) | 100% |
| 3 | 23 | 26.12 (0.62) | 28.88 (0.90) | 32.10 (1.03) | 29.36 (1.35) | 100% |
| 1 | 23 | 26.02 (0.74) | 29.50 (1.77) | 32.90 (0.57) | 29.83 (2.09) | 100% |
| 0.5 | 3 | 26.00 (1.43) | 31.01 (0.64) | 31.99 (0.34) | 30.60 (1.76) | 100% |

**Table 2.** Confirmation of the LOD finding study (1 viral RNA copy per µl) in an individual sample and a pooled sample.

| Viral<br>RNA<br>Copy/$\mu\text{l}$ | N | Pool<br>size | Mean Ct values (SD) | | | | %<br>Triggerin<br>g Reflex |
| --- | --- | --- | --- | --- | --- | --- | --- |
|  |  |  | MS2 Phage | N gene | ORF1ab | S gene |  |
| 1 | 3 | 1 | 28.81 (0.63) | 28.20 (0.10) | 30.20 (0.31) | 29.50 (0.34) | N/A |
|  | 9 | 24 | 27.55 (0.61) | 31.11 (0.29) | 33.19 (0.51) | 32.60 (0.38) | 100% |
| 6 | 3 | 1 | 26.35(0.54) | 27.77(1.11) | 29.58(0.24) | 27.26(0.41) | N/A |
|  | 20 | 24 | 28.55(0.62) | 30.33(0.91) | 33.33(0.39) | 30.11(1.7) | 100% |

During the 20-week period from August 27, 2020 to January 13, 2021, 249,531 specimens condensed into 10,057 pools were collected from K-12 students and faculty and staff at 93 schools (Supplementary Table 2). An additional 15,802 specimens condensed into 715 pools were collected from 18 universities (Supplementary Table 3). The number of pools collected from each of the 111 institutions ranged from 1 to 92 pools (mean=13, median=7) per school. Each pool contained 2 to 24 (mean=21, median=23) individual saliva samples. Of the 10,772 pools, 371 pools were positive (3.4% pool positivity rate), with 1 to 14 (mean=0.8, median=0) positive pools per institution. A total of 863 positive individual samples (0.2% individual positivity rate) were detected using this method. Each positive pool had 1 to 24 (mean=1.4, median=0) samples that tested positive on deconvolution testing.

The results of pooled testing were available and reported to the institutions within 6-12 hours from specimen accessioning on average and within 24 hours from specimen collection. The individual results were reported within 12-24 hours from specimen accessioning on average and within 36-48 hours from specimen collection. The cost of the initial pooled testing was approximately $13-15 per individual for all participating schools. Reflex testing included additional charges, with the overall pooled to individual test averaging $16-17 per person. This included all testing supplies, logistics, and personnel as well as diagnostic reporting, and allowed for over a dozen districts to participate in the current study.

In analyzing the trend of positivity among the pools (Figure 1), observable peaks were seen on days following long weekends and holidays. The highest number of positive pools was observed 5 days after New Year’s Eve (January 5, 2021; 39 positive pools, 230 were positive individuals). This was a significant increase in the number of positive samples (p=0.045; Figure 2A) compared to any other testing day. The second highest number of positive pools was recorded on November 11 (29 positive pools; 32 positive individuals) and the second highest number of positive individual samples was observed on November 13 (28 positive pools; 50 positive individuals), 11 to 13 days after Halloween. This was also a significant increase when compared to the number of positive samples from prior dates (p=0.0048; Figure 2B). Of note, the SalivaClear™ method was able to identify 10 positive (out of 16) pools for one school campus on November 13, corresponding to 26 positive individuals who attended an off-campus Halloween celebration. The saliva pooling method also identified 10 positive (out of 64) pools from another school on November 10, and all positive individuals from these pools had reported separate exposures. The third highest number of positive pools was on December 1 and December 3 (21 positive pools each; 26 and 22 positive individuals, respectively), 5 to 7 days after Thanksgiving; however, this increase was not statistically significant (p=0.600).

**Figure 1.**
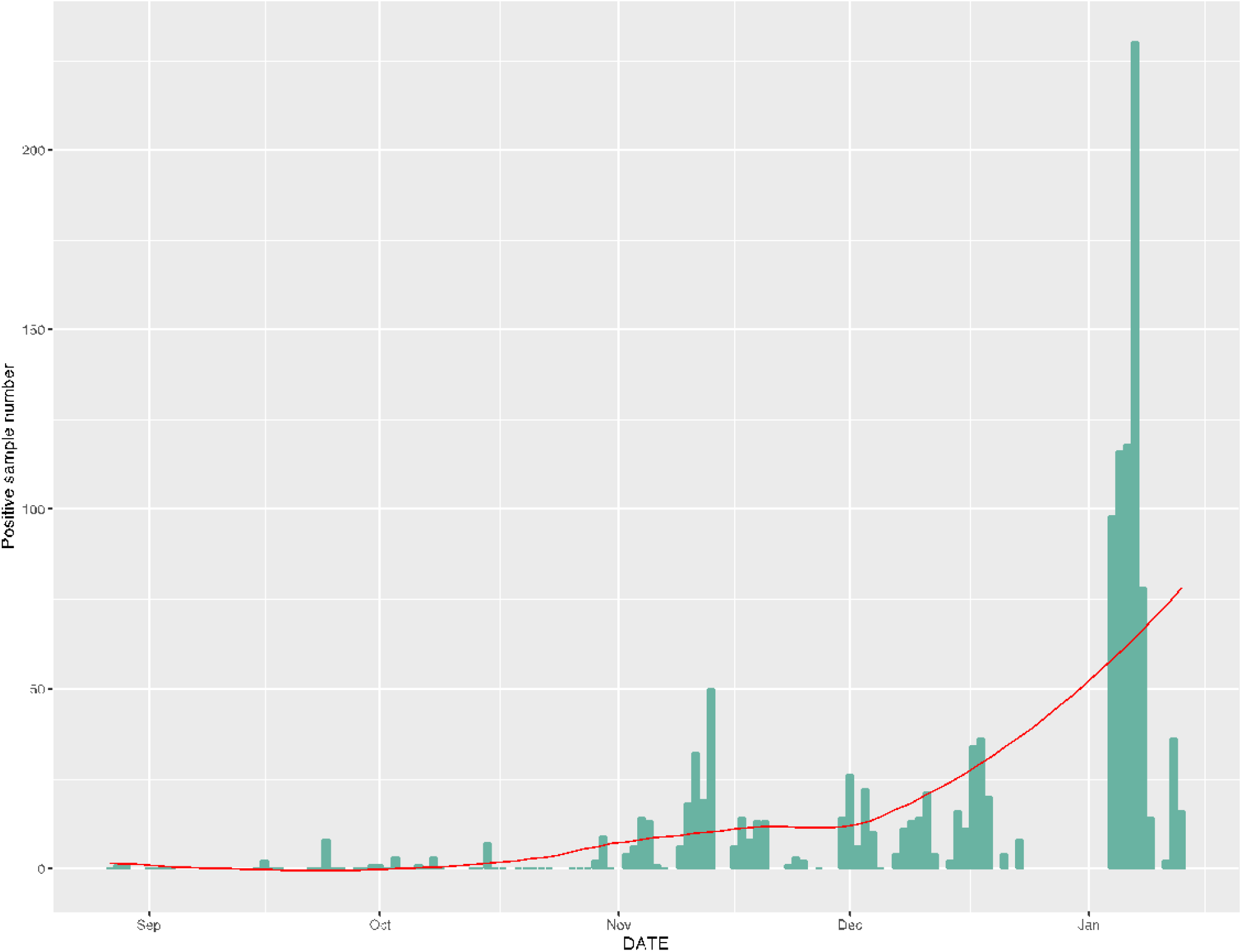
Trend of saliva-based pooled testing positivity in 109 K-12 schools and universities for a five-month period. Peaks of positive cases (pooled samples) were observed in dates following Halloween and New Year’s Eve.

**Figure 2.**
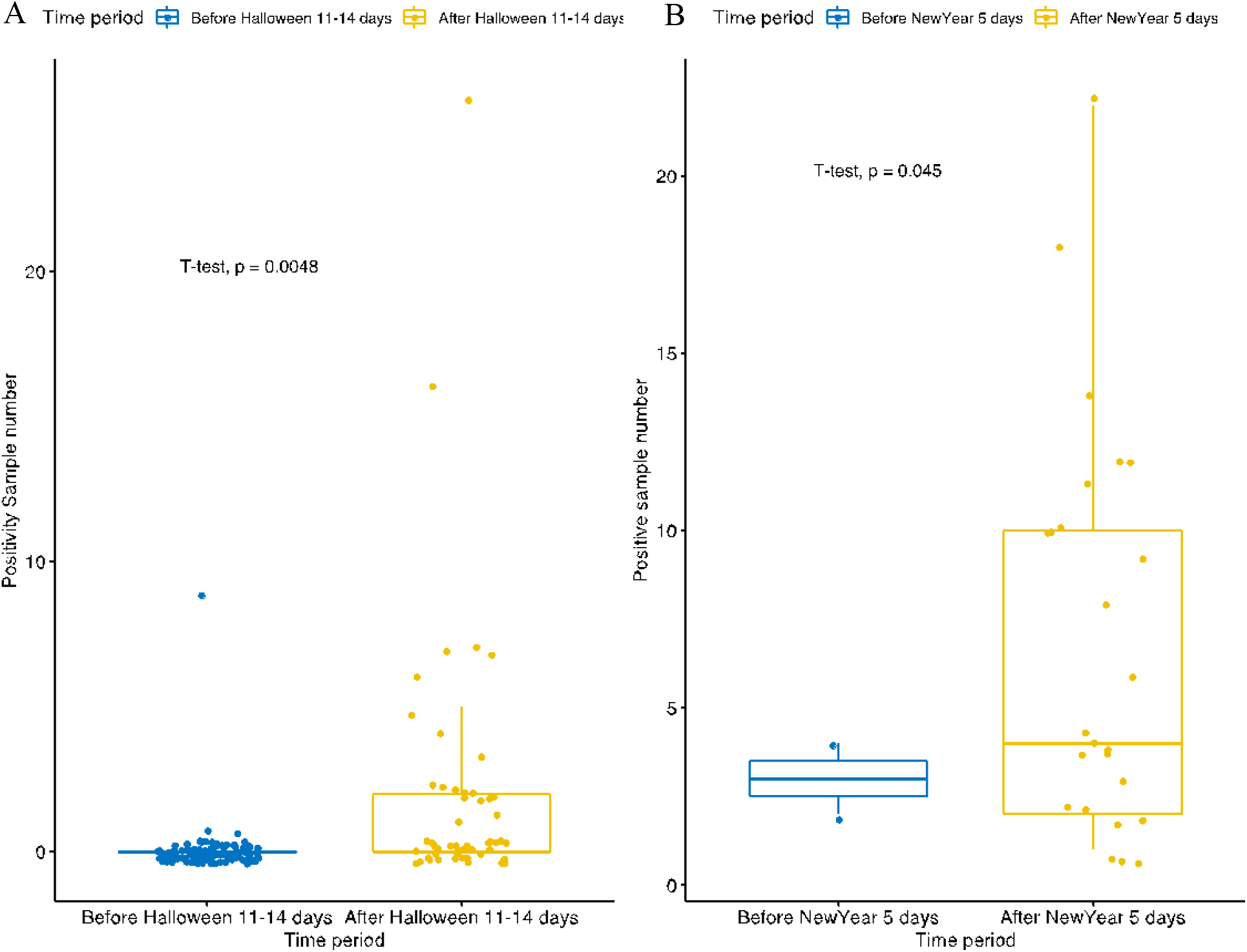
Box-and-whisker scatter plot of positive cases detected 7-8 days after New Year’s Eve (panel A) and 11-14 days after Halloween (panel B). The upper and lower sides of the box correspond to the 75^th^ and 25^th^ percentiles, respectively. The ends of the whiskers correspond to the highest and lowest value. Each dot indicates the number of positive cases in one organization.

From the pools obtained through SalivaClear™ testing, we randomly selected 20 positive pools and compared the CT values of the pool to the corresponding positive individual sample in that pool. There was a 100% agreement in the detection of viral targets between the pools and individual samples (Figure 3; Supplementary Table 4). The average difference in CT values between the individual sample and the pooled sample were 2.10, 2.97 and 3.23 for the three targets (N gene, S gene, Orf1ab gene), respectively. Approximately 2% of all negative pools were deconvoluted to check if some positive individual samples may be missed by pooling. All individual samples also tested negative (data not shown).

**Figure 3.**
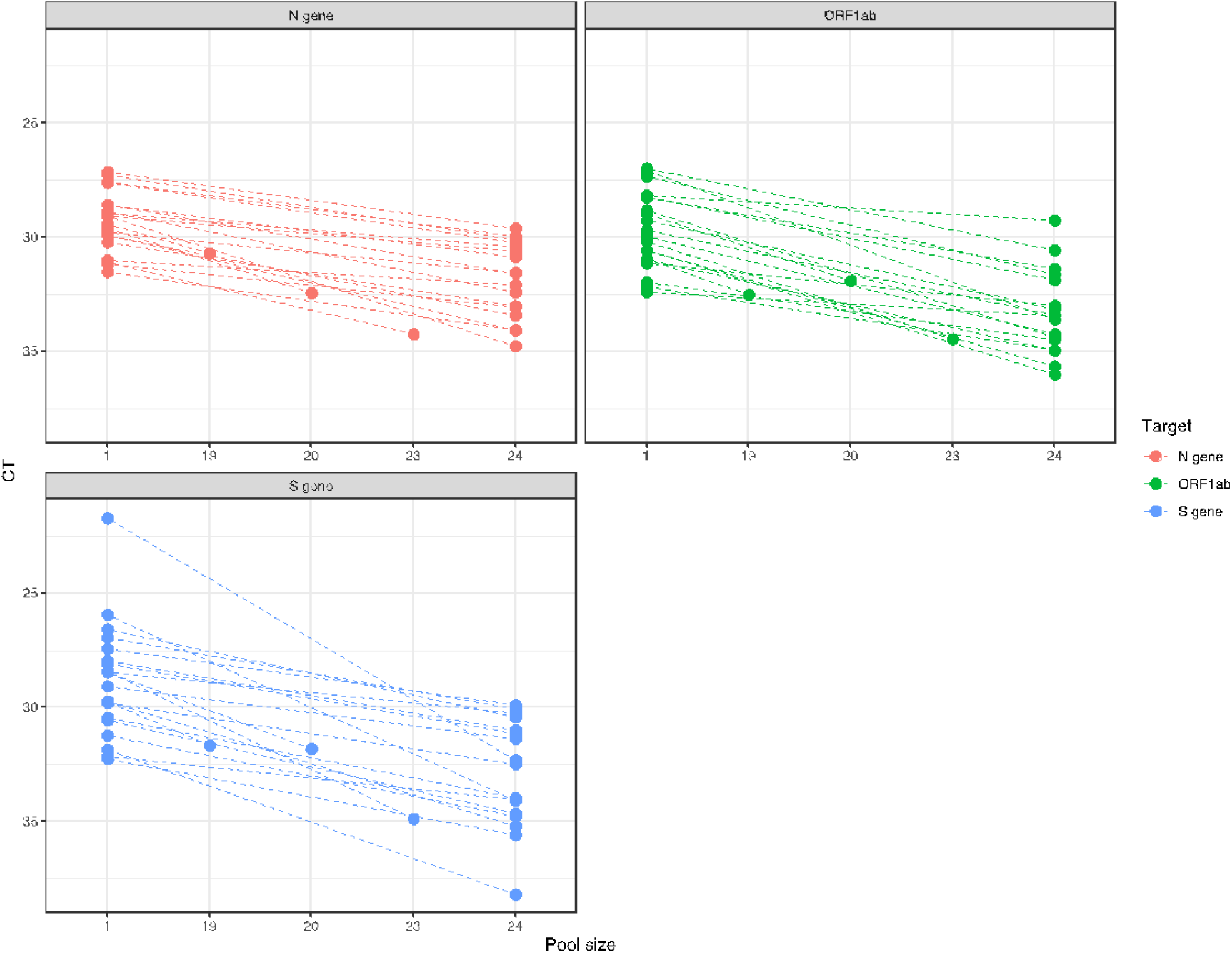
CT values comparisons between pooled and individual samples. Twenty randomly selected positive samples were analyzed for comparison of CT values when detected in a pool of 24 samples and when analyzed as a single sample. The dashed lines connect the dots that indicate the CT values of individual samples (higher) to the corresponding pools (lower).

## Discussion

The closure of schools has been one of the most adverse impacts of Covid-19 among children^15^. Other than negatively affecting learning aptitude especially in younger children, school closures have been linked to several psychosocial problems^16^. The full negative impact of school closures due to COVID-19 throughout the world will remain unknown for years to come; however, the effects on mental health, nutrition, increasing educational gaps and socioeconomical inequality have already become apparent^15^. In the 5^th^ largest school district in the nation, schools are being forced to reopen despite continuously high positivity rates due to a surge student suicide^17^, demonstrating how important schools are for meeting both academic and nonacademic needs of our nation’s children.

To meet the COVID-19 testing demands needed for schools to re-open, we sought to create an organizational-based pooling strategy, whereby schools could enroll in surveillance testing to monitor groups of people through pooled testing. By using organizational-based pooling, the program avoided random pooling of samples. Instead, our approach pooled groups of known contacts from school where students, faculty and staff regularly interacted utilizing a similar protocol. By this approach, a positive case impacted the entire pooled population, and the entire pooled population was treated as an infected cohort until further individual testing was completed and reported through a healthcare provider.

Here, we report on testing of over 400,000 Covid-19 RT-PCR specimens from students, faculty and staff from 93 K-12 schools and 18 universities during a 20-week period during this pandemic. The pooled testing was facilitated by trained collection specialists and utilized local logistics companies, pre-barcoded tubes, and automation to expedite processing. We expect this passive drool method minimized the potential aerosolization of infectious agents as seen in involuntary coughing and sneezing encountered with the swab methods (nasopharyngeal, oropharyngeal, nasal). These factors enabled the collection and assessment of up to 25,000 tests per day, highlighting the potential to scale such a method.

Importantly, we found that SalivaClear™ has a similar sensitivity to the molecular assay of individual samples, in terms of both qualitative (100% agreement of results on both pooled and individual samples) and quantitative (comparable CT values between pooled and individual samples) measures. Without sacrificing the reliability of the molecular assay, pooling of samples had substantially reduced the costs associated with PCR testing. In order to broadly test communities and schools, lower pricing and pooled approaches in particular allow for a “multiplier effect” that can provide significant economies of scale, potentially decreasing the costs even further at larger scale.

The rapid turn-around time of pooled results allowed schools to assess transmission and adjust prevention protocols in a timely manner. As a result of sampling at the same time, coupled with grouping by work area, grade, or section, pool results helped institution administrators to determine if transmission has occurred, make data driven decisions, and adjust and improve safety protocols. The surveillance program is just one component of the established protocols essential for preventing transmission of Covid-19 within the school premises and limiting outbreaks. In one instance, in-school transmission of the virus was determined within the main office and led to a review of heating, ventilating and air-conditioning (HVAC) systems as well as air circulation mapping throughout the workstations. Surprisingly, although plexiglass dividers were installed to prevent viral transmission, the air circulation testing using smoke devices revealed that plexiglass dividers, when coupled with side panels, significantly impeded air circulation leading to increased viral transmission.

Several studies have shown reliable detection of SARS-CoV-2 RNA in saliva samples^18-23^. In a few of them, saliva samples had superior sensitivity and stability compared to the nasopharyngeal swab^18,22,24^. Other than causing significant exposure to healthcare workers and discomfort to patients, nasopharyngeal swabs were limited by finite supply chains and restricted distribution. Our data also demonstrated adequate stability of saliva samples when collected in dry, sterile collection containers, thereby expanding the options for Covid-19 specimen collection device (Supplemental Table 5). Additionally, saliva collection is less invasive and can be self-collected under minimal supervision of a healthcare provider.

Here we describe the pooled method of SalivaClear™ that uses a large pool size that has not previously been described in the literature, enabling a greater “multiplier” effect to increase testing capacity across the county. With testing result delays across the country, primarily due to reagent and capacity shortages, pooling enables conversation of resources while scaling capacity and still being able to provide individualized results when required. Our data shows that pooled saliva testing performed with the highest efficiency in terms of timeliness and cost may be one of the solutions in supporting school organizations to safely conduct in-person classes amidst the pandemic crisis.

One limitation of this study is that the applicability of the methodology described here requires both technical and logistical methods that are not reproducible in every lab. However, concepts of pooling, strategies around reducing costs for schools, and the unique value of providing testing for schools as it relates to community viral spread described here, remain broadly applicable to the K-12 testing strategies that are required around the country. Additional limitations include the need for lab-based testing as in theory this concept can be broadly applied to point of care concepts as well. However, as with any testing strategy, scalability is important. The ability to rapidly scale SalivaClear™ using existing lab infrastructure makes it broadly appealing to labs seeking to increase testing capacity and efficiency. While the non-standardized methods of school openings across the country are a limiting factor when evaluating the data for the exact role SalivaClear™ played for each school, it is important to highlight that all schools utilizing this method were able to remain open.

In order to provide enough testing capacity to safely open schools and provide testing for at-risk communities, pooled concepts must be further evaluated and considered. As vaccination against COVID-19 may remain unavailable for children under 16 years of age for months to come, pooled testing must be considered a fundamental component to the safe reopening of schools while minimizing transmission among students and faculty^23^.

## Data Availability

The raw data set for this study is available on a secure server hosted by Mirimus Clinical Labs, Brooklyn, NY.

## Conflict of interest

None

## Funding

Skoll Foundation generously provided funding to Mobilizing Foundation and Mirimus for these studies.

## Notes

### Competing Interest Statement

The authors have declared no competing interest.

### Author Declarations

The SUNY Downstate Health Sciences University Institutional Review Board (IRB) reviewed and approved the study protocol (IRB #1232938-3).

