## Supplementary Table 1 for "Pooled Surveillance Testing Program for Asymptomatic SARS-CoV-2 Infections in K-12 Schools and Universities"

Supplementary Table 1. Assessment of pooled specimens with 1-12 viral copies per microliter of specimen, in pool sizes ranging from 1 to 24. To generate each pool, 200µL of the known positive stock solution of 1-12 copies/µL of specimen was combined with known negative saliva matrix. For each RNA extraction, 200µL of each sample was used.

| **Viral RNA Copy/µL** | **N** | **Pool size** | Mean Ct values (SD; N=3) | | | | **% Triggering Reflex** |
|---|---|---|---|---|---|---|---|
| | | | **MS2** | **N gene** | **Orf1ab** | **S gene** | |
| 12 copies | 3 | 1 | 28.83 (0.57) | 25.4 (0.3) | 26.81 (0.38) | 26.01 (0.5) | N/A |
| | 3 | 8 | 29.63 (0.24) | 28.42 (0.32) | 30.24 (0.33) | 29.03 (0.24) | 100% |
| | 3 | 16 | 28.72 (0.34) | 28.52 (0.19) | 30.22 (0.1) | 29.32 (0.13) | 100% |
| | 9 | 24 | 28.68 (1.28) | 29.08 (0.95) | 31.54 (0.66) | 29.96 (1.02) | 100% |
| | 3 | 36 | 28.41 (0.23) | 30.51 (0.56) | 32.5 (0.69) | 32 (0.67) | 100% |
| | 3 | 48 | 29.91 (0.33) | 30.91 (0.42) | 33.3 (0.28) | 32 (0.27) | 100% |
| 6 copies | 3 | 1 | 28.24 (0.45) | 25.85 (0.32) | 27.74 (0.18) | 27.74 (0.77) | N/A |
| | 3 | 8 | 28.11 (0.21) | 29.31 (0.68) | 31.1 (0.65) | 30.11 (0.8) | 100% |
| | 3 | 16 | 28.05 (0.44) | 29.45 (0.38) | 31.35 (0.24) | 29.95 (0.64) | 100% |
| | 9 | 24 | 29.1 (0.6) | 30.39 (0.68) | 32.87 (0.97) | 31.78 (0.68) | 100% |
| | 3 | 36 | 28.59 (0.46) | 31.96 (0.28) | 34.23 (0.34) | 32.75 (0.72) | 100% |
| | 3 | 48 | 28.84 (0.34) | 31.79 (0.61) | 33.56 (0.97) | 31.59 (0.29) | 100% |
| 3 copies | 3 | 1 | 27.54 (0.66) | 27.15 (0.94) | 28.92 (0.73) | 27.74 (0.88) | N/A |
| | 3 | 8 | 27.35 (0.44) | 29.21 (0.87) | 32.74 (0.23) | 30.88 (2.43) | 100% |
| | 3 | 16 | 27.83 (0.08) | 30.5 (1.24) | 32.97 (0.02) | 32.08 (0.71) | 100% |
| | 9 | 24 | 28.17 (0.41) | 30.68 (0.47) | 32.99 (0.99) | 32.08 (0.8) | 100% |
| | 3 | 36 | 28.88 (0.34) | 31.82 (0.22) | 34.18 (NA) | 31.72 (NA) | 100% |
| | 3 | 48 | 28.02 (0.3) | 31.1 (0.18) | 34.38 (0.21) | 33.58 (0.27) | 100% |
| 1 copy | 3 | 1 | 28.81 (0.63) | 28.2 (0.1) | 30.2 (0.31) | 29.5 (0.34) | N/A |
| | 3 | 8 | 28.87 (0.69) | 30.15 (1.17) | 33.3 (0.57) | 31.24 (0.73) | 100% |
| | 3 | 16 | 28.36 (0.7) | 29.52 (0.76) | 33.67 (0.9) | 31.83 (0.62) | 100% |
| | 9 | 24 | 27.55 (0.61) | 31.11 (0.29) | 33.19 (0.51) | 32.6 (0.38) | 100% |
| | 3 | 36 | 28.73 (0.62) | 31.72 (0.81) | 34.11 (0.09) | 32.12 (1.46) | 100% |
| | 3 | 48 | 28.61 (0.47) | 31.61 (0.61) | 34.22 (0.18) | 33.62 (2.05) | 100% |
