## Supplementary Table 2 for "Pooled Surveillance Testing Program for Asymptomatic SARS-CoV-2 Infections in K-12 Schools and Universities"

Supplementary Table 2. Raw data of each K-12 organization tested between Aug 27, 2020 and Jan 13, 2021.

| K-12 NO. | Pool number | Pool size | Pool Positive number | Pool Positivity rate | Individual number | Individual Positive number | Individual Positivity rate | Date |
|---|---|---|---|---|---|---|---|---|
| R324851 | 8 | 24 | 0 | 0.00% | 192 | 0 | 0.00% | 8/27/20 |
| R324833 | 75 | 23 | 1 | 1.33% | 1711 | 1 | 0.06% | 8/28/20 |
| R324833 | 10 | 21 | 1 | 10.00% | 209 | 1 | 0.48% | 8/29/20 |
| R324851 | 6 | 25 | 0 | 0.00% | 147 | 0 | 0.00% | 8/29/20 |
| R324855 | 13 | 21 | 0 | 0.00% | 272 | 0 | 0.00% | 9/2/20 |
| R324851 | 10 | 22 | 0 | 0.00% | 223 | 0 | 0.00% | 9/2/20 |
| R326854 | 2 | 23 | 0 | 0.00% | 45 | 0 | 0.00% | 9/3/20 |
| R324833 | 46 | 22 | 0 | 0.00% | 1026 | 0 | 0.00% | 9/4/20 |
| R324851 | 1 | 11 | 0 | 0.00% | 11 | 0 | 0.00% | 9/4/20 |
| R326854 | 3 | 19 | 0 | 0.00% | 56 | 0 | 0.00% | 9/4/20 |
| R326820 | 3 | 18 | 0 | 0.00% | 53 | 0 | 0.00% | 9/9/20 |
| R324851 | 1 | 10 | 0 | 0.00% | 10 | 0 | 0.00% | 9/9/20 |
| R324836 | 49 | 22 | 0 | 0.00% | 1095 | 0 | 0.00% | 9/10/20 |
| R324833 | 47 | 23 | 0 | 0.00% | 1070 | 0 | 0.00% | 9/11/20 |
| R325241 | 4 | 23 | 0 | 0.00% | 93 | 0 | 0.00% | 9/16/20 |
| R324836 | 50 | 23 | 0 | 0.00% | 1145 | 0 | 0.00% | 9/17/20 |
| R324833 | 48 | 23 | 0 | 0.00% | 1083 | 0 | 0.00% | 9/18/20 |
| R325305 | 1 | 22 | 0 | 0.00% | 22 | 0 | 0.00% | 9/18/20 |
| R324851 | 5 | 25 | 0 | 0.00% | 126 | 0 | 0.00% | 9/18/20 |
| R326854 | 1 | 17 | 0 | 0.00% | 17 | 0 | 0.00% | 9/18/20 |
| R325241 | 43 | 24 | 0 | 0.00% | 1030 | 0 | 0.00% | 9/22/20 |
| R326820 | 4 | 24 | 0 | 0.00% | 96 | 0 | 0.00% | 9/23/20 |
| R325249 | 3 | 23 | 0 | 0.00% | 69 | 0 | 0.00% | 9/24/20 |
| R325260 | 2 | 20 | 0 | 0.00% | 40 | 0 | 0.00% | 9/24/20 |
| R324836 | 54 | 21 | 0 | 0.00% | 1123 | 0 | 0.00% | 9/24/20 |
| R324833 | 46 | 24 | 0 | 0.00% | 1108 | 0 | 0.00% | 9/25/20 |
| R324851 | 6 | 24 | 0 | 0.00% | 142 | 0 | 0.00% | 9/25/20 |
| R326854 | 2 | 19 | 0 | 0.00% | 37 | 0 | 0.00% | 9/25/20 |
| R325249 | 2 | 20 | 0 | 0.00% | 40 | 0 | 0.00% | 9/28/20 |
| R325241 | 45 | 23 | 0 | 0.00% | 1034 | 0 | 0.00% | 9/29/20 |
| R325249 | 7 | 21 | 0 | 0.00% | 146 | 0 | 0.00% | 10/1/20 |
| R324836 | 56 | 20 | 0 | 0.00% | 1141 | 0 | 0.00% | 10/1/20 |
| R325249 | 8 | 16 | 0 | 0.00% | 129 | 0 | 0.00% | 10/2/20 |
| R324833 | 46 | 24 | 0 | 0.00% | 1093 | 0 | 0.00% | 10/2/20 |
| R325136 | 5 | 24 | 0 | 0.00% | 120 | 0 | 0.00% | 10/2/20 |
| R324851 | 7 | 22 | 0 | 0.00% | 154 | 0 | 0.00% | 10/2/20 |
| R326854 | 1 | 22 | 0 | 0.00% | 22 | 0 | 0.00% | 10/2/20 |
| R325137 | 32 | 24 | 3 | 9.38% | 768 | 1 | 0.13% | 10/3/20 |
| R326820 | 7 | 18 | 1 | 14.29% | 128 | 1 | 0.78% | 10/6/20 |
| R325249 | 6 | 18 | 0 | 0.00% | 105 | 0 | 0.00% | 10/6/20 |
| R325241 | 46 | 22 | 0 | 0.00% | 1025 | 0 | 0.00% | 10/6/20 |
| R325136 | 2 | 24 | 0 | 0.00% | 48 | 0 | 0.00% | 10/7/20 |
| R325139 | 1 | 23 | 0 | 0.00% | 23 | 0 | 0.00% | 10/7/20 |
| R325305 | 60 | 24 | 2 | 3.33% | 1416 | 2 | 0.14% | 10/8/20 |
| R325147 | 34 | 23 | 1 | 2.94% | 795 | 1 | 0.13% | 10/8/20 |
| R326820 | 5 | 23 | 0 | 0.00% | 117 | 0 | 0.00% | 10/8/20 |
| R325136 | 3 | 24 | 0 | 0.00% | 72 | 0 | 0.00% | 10/8/20 |
| R324836 | 56 | 20 | 0 | 0.00% | 1110 | 0 | 0.00% | 10/8/20 |
| R325125 | 3 | 24 | 0 | 0.00% | 72 | 0 | 0.00% | 10/8/20 |
| R326820 | 2 | 26 | 0 | 0.00% | 51 | 0 | 0.00% | 10/9/20 |
| R325249 | 12 | 23 | 0 | 0.00% | 280 | 0 | 0.00% | 10/9/20 |
| R324833 | 53 | 21 | 0 | 0.00% | 1093 | 0 | 0.00% | 10/9/20 |

| | | | | | | | | |
|---|---|---|---|---|---|---|---|---|
| R325137 | 25 | 24 | 0 | 0.00% | 588 | 0 | 0.00% | 10/9/20 |
| R325148 | 6 | 23 | 0 | 0.00% | 139 | 0 | 0.00% | 10/9/20 |
| R324851 | 7 | 24 | 0 | 0.00% | 169 | 0 | 0.00% | 10/9/20 |
| R326854 | 4 | 22 | 0 | 0.00% | 89 | 0 | 0.00% | 10/9/20 |
| R325125 | 5 | 21 | 0 | 0.00% | 106 | 0 | 0.00% | 10/9/20 |
| R325134 | 4 | 23 | 0 | 0.00% | 91 | 0 | 0.00% | 10/13/20 |
| R325241 | 47 | 22 | 0 | 0.00% | 1033 | 0 | 0.00% | 10/13/20 |
| R325129 | 3 | 22 | 0 | 0.00% | 65 | 0 | 0.00% | 10/13/20 |
| R325137 | 1 | 18 | 0 | 0.00% | 18 | 0 | 0.00% | 10/13/20 |
| R325138 | 33 | 24 | 0 | 0.00% | 779 | 0 | 0.00% | 10/13/20 |
| R325134 | 11 | 24 | 0 | 0.00% | 261 | 0 | 0.00% | 10/14/20 |
| R325136 | 2 | 24 | 0 | 0.00% | 48 | 0 | 0.00% | 10/14/20 |
| R325145 | 2 | 22 | 0 | 0.00% | 43 | 0 | 0.00% | 10/14/20 |
| R325125 | 11 | 24 | 1 | 9.09% | 264 | 1 | 0.38% | 10/15/20 |
| R324836 | 56 | 20 | 1 | 1.79% | 1100 | 6 | 0.55% | 10/15/20 |
| R325155 | 9 | 17 | 0 | 0.00% | 156 | 0 | 0.00% | 10/15/20 |
| R325128 | 6 | 23 | 0 | 0.00% | 140 | 0 | 0.00% | 10/15/20 |
| R325755 | 4 | 23 | 0 | 0.00% | 91 | 0 | 0.00% | 10/15/20 |
| R325127 | 4 | 24 | 0 | 0.00% | 96 | 0 | 0.00% | 10/15/20 |
| R325151 | 12 | 24 | 0 | 0.00% | 285 | 0 | 0.00% | 10/15/20 |
| R325147 | 14 | 23 | 0 | 0.00% | 323 | 0 | 0.00% | 10/16/20 |
| R325249 | 14 | 24 | 0 | 0.00% | 329 | 0 | 0.00% | 10/16/20 |
| R324833 | 53 | 21 | 0 | 0.00% | 1098 | 0 | 0.00% | 10/16/20 |
| R342986 | 3 | 21 | 0 | 0.00% | 64 | 0 | 0.00% | 10/16/20 |
| R325129 | 3 | 24 | 0 | 0.00% | 72 | 0 | 0.00% | 10/16/20 |
| R325137 | 32 | 24 | 0 | 0.00% | 761 | 0 | 0.00% | 10/16/20 |
| R325305 | 65 | 22 | 0 | 0.00% | 1433 | 0 | 0.00% | 10/16/20 |
| R324851 | 8 | 23 | 0 | 0.00% | 185 | 0 | 0.00% | 10/16/20 |
| R326854 | 4 | 21 | 0 | 0.00% | 83 | 0 | 0.00% | 10/16/20 |
| R325125 | 20 | 24 | 0 | 0.00% | 480 | 0 | 0.00% | 10/16/20 |
| R325136 | 3 | 24 | 0 | 0.00% | 72 | 0 | 0.00% | 10/17/20 |
| R326820 | 13 | 24 | 0 | 0.00% | 316 | 0 | 0.00% | 10/19/20 |
| R325554 | 3 | 24 | 0 | 0.00% | 72 | 0 | 0.00% | 10/19/20 |
| R325249 | 9 | 22 | 0 | 0.00% | 200 | 0 | 0.00% | 10/19/20 |
| R334362 | 37 | 21 | 0 | 0.00% | 795 | 0 | 0.00% | 10/19/20 |
| R325755 | 1 | 12 | 0 | 0.00% | 12 | 0 | 0.00% | 10/19/20 |
| R324727 | 15 | 24 | 0 | 0.00% | 354 | 0 | 0.00% | 10/19/20 |
| R325127 | 4 | 19 | 0 | 0.00% | 76 | 0 | 0.00% | 10/19/20 |
| R326854 | 7 | 21 | 0 | 0.00% | 147 | 0 | 0.00% | 10/19/20 |
| R325134 | 10 | 22 | 0 | 0.00% | 224 | 0 | 0.00% | 10/20/20 |
| R325241 | 46 | 23 | 0 | 0.00% | 1038 | 0 | 0.00% | 10/20/20 |
| R325127 | 5 | 24 | 0 | 0.00% | 118 | 0 | 0.00% | 10/20/20 |
| R325138 | 25 | 23 | 0 | 0.00% | 585 | 0 | 0.00% | 10/20/20 |
| R339851 | 10 | 24 | 0 | 0.00% | 235 | 0 | 0.00% | 10/20/20 |
| R334327 | 5 | 24 | 0 | 0.00% | 120 | 0 | 0.00% | 10/20/20 |
| R325125 | 37 | 24 | 0 | 0.00% | 870 | 0 | 0.00% | 10/20/20 |
| R425408 | 16 | 23 | 0 | 0.00% | 363 | 0 | 0.00% | 10/20/20 |
| R325147 | 10 | 23 | 0 | 0.00% | 228 | 0 | 0.00% | 10/21/20 |
| R339853 | 9 | 25 | 0 | 0.00% | 223 | 0 | 0.00% | 10/21/20 |
| R324855 | 10 | 24 | 0 | 0.00% | 237 | 0 | 0.00% | 10/21/20 |
| R334335 | 5 | 16 | 0 | 0.00% | 81 | 0 | 0.00% | 10/21/20 |
| R325136 | 2 | 24 | 0 | 0.00% | 48 | 0 | 0.00% | 10/21/20 |
| R325127 | 5 | 21 | 0 | 0.00% | 104 | 0 | 0.00% | 10/21/20 |
| R325529 | 3 | 24 | 0 | 0.00% | 71 | 0 | 0.00% | 10/21/20 |
| R325125 | 10 | 24 | 0 | 0.00% | 240 | 0 | 0.00% | 10/21/20 |
| R334324 | 3 | 18 | 0 | 0.00% | 53 | 0 | 0.00% | 10/21/20 |
| R325155 | 8 | 24 | 0 | 0.00% | 192 | 0 | 0.00% | 10/22/20 |
| R325147 | 28 | 18 | 0 | 0.00% | 510 | 0 | 0.00% | 10/22/20 |

| | | | | | | | | |
|---|---|---|---|---|---|---|---|---|
| R325134 | 6 | 23 | 0 | 0.00% | 139 | 0 | 0.00% | 10/22/20 |
| R339853 | 7 | 21 | 0 | 0.00% | 150 | 0 | 0.00% | 10/22/20 |
| R325249 | 15 | 23 | 0 | 0.00% | 346 | 0 | 0.00% | 10/22/20 |
| R325128 | 11 | 24 | 0 | 0.00% | 259 | 0 | 0.00% | 10/22/20 |
| R325755 | 6 | 24 | 0 | 0.00% | 145 | 0 | 0.00% | 10/22/20 |
| R325136 | 3 | 24 | 0 | 0.00% | 72 | 0 | 0.00% | 10/22/20 |
| R324836 | 56 | 20 | 0 | 0.00% | 1113 | 0 | 0.00% | 10/22/20 |
| R325151 | 20 | 24 | 0 | 0.00% | 480 | 0 | 0.00% | 10/22/20 |
| R325125 | 10 | 24 | 0 | 0.00% | 240 | 0 | 0.00% | 10/22/20 |
| R325147 | 12 | 24 | 0 | 0.00% | 287 | 0 | 0.00% | 10/23/20 |
| R339853 | 8 | 24 | 0 | 0.00% | 194 | 0 | 0.00% | 10/23/20 |
| R324833 | 55 | 20 | 0 | 0.00% | 1099 | 0 | 0.00% | 10/23/20 |
| R334362 | 1 | 7 | 0 | 0.00% | 7 | 0 | 0.00% | 10/23/20 |
| R342986 | 5 | 19 | 0 | 0.00% | 93 | 0 | 0.00% | 10/23/20 |
| R325129 | 4 | 23 | 0 | 0.00% | 91 | 0 | 0.00% | 10/23/20 |
| R325137 | 25 | 23 | 0 | 0.00% | 586 | 0 | 0.00% | 10/23/20 |
| R325305 | 58 | 25 | 0 | 0.00% | 1423 | 0 | 0.00% | 10/23/20 |
| R324851 | 8 | 23 | 0 | 0.00% | 187 | 0 | 0.00% | 10/23/20 |
| R326854 | 3 | 25 | 0 | 0.00% | 75 | 0 | 0.00% | 10/23/20 |
| R325125 | 7 | 24 | 0 | 0.00% | 167 | 0 | 0.00% | 10/23/20 |
| R339853 | 5 | 24 | 0 | 0.00% | 121 | 0 | 0.00% | 10/26/20 |
| R325249 | 12 | 21 | 0 | 0.00% | 249 | 0 | 0.00% | 10/26/20 |
| R324727 | 7 | 23 | 0 | 0.00% | 162 | 0 | 0.00% | 10/26/20 |
| R325134 | 10 | 23 | 0 | 0.00% | 225 | 0 | 0.00% | 10/27/20 |
| R325241 | 46 | 23 | 0 | 0.00% | 1037 | 0 | 0.00% | 10/27/20 |
| R325127 | 7 | 22 | 0 | 0.00% | 151 | 0 | 0.00% | 10/27/20 |
| R325138 | 33 | 24 | 0 | 0.00% | 779 | 0 | 0.00% | 10/27/20 |
| R339851 | 38 | 23 | 0 | 0.00% | 892 | 0 | 0.00% | 10/27/20 |
| R325125 | 40 | 24 | 0 | 0.00% | 960 | 0 | 0.00% | 10/27/20 |
| R425408 | 30 | 24 | 0 | 0.00% | 720 | 0 | 0.00% | 10/27/20 |
| R325147 | 15 | 22 | 0 | 0.00% | 333 | 0 | 0.00% | 10/28/20 |
| R339853 | 4 | 24 | 0 | 0.00% | 97 | 0 | 0.00% | 10/28/20 |
| R334335 | 5 | 18 | 0 | 0.00% | 88 | 0 | 0.00% | 10/28/20 |
| R334362 | 36 | 21 | 0 | 0.00% | 773 | 0 | 0.00% | 10/28/20 |
| R325129 | 5 | 22 | 0 | 0.00% | 111 | 0 | 0.00% | 10/28/20 |
| R334342 | 4 | 23 | 0 | 0.00% | 93 | 0 | 0.00% | 10/28/20 |
| R325148 | 6 | 21 | 0 | 0.00% | 126 | 0 | 0.00% | 10/28/20 |
| R325529 | 7 | 22 | 0 | 0.00% | 152 | 0 | 0.00% | 10/28/20 |
| R334327 | 5 | 23 | 0 | 0.00% | 117 | 0 | 0.00% | 10/28/20 |
| R325125 | 32 | 24 | 0 | 0.00% | 759 | 0 | 0.00% | 10/28/20 |
| R334324 | 3 | 19 | 0 | 0.00% | 56 | 0 | 0.00% | 10/28/20 |
| R325249 | 16 | 23 | 3 | 18.75% | 371 | 1 | 0.27% | 10/29/20 |
| R325305 | 59 | 24 | 2 | 3.39% | 1417 | 1 | 0.07% | 10/29/20 |
| R325155 | 8 | 24 | 0 | 0.00% | 188 | 0 | 0.00% | 10/29/20 |
| R325147 | 18 | 21 | 0 | 0.00% | 386 | 0 | 0.00% | 10/29/20 |
| R325134 | 6 | 24 | 0 | 0.00% | 142 | 0 | 0.00% | 10/29/20 |
| R325128 | 12 | 24 | 0 | 0.00% | 292 | 0 | 0.00% | 10/29/20 |
| R325755 | 7 | 22 | 0 | 0.00% | 156 | 0 | 0.00% | 10/29/20 |
| R325136 | 2 | 24 | 0 | 0.00% | 48 | 0 | 0.00% | 10/29/20 |
| R342986 | 1 | 20 | 0 | 0.00% | 20 | 0 | 0.00% | 10/29/20 |
| R324836 | 56 | 20 | 0 | 0.00% | 1122 | 0 | 0.00% | 10/29/20 |
| R325145 | 2 | 24 | 0 | 0.00% | 47 | 0 | 0.00% | 10/29/20 |
| R325151 | 20 | 24 | 0 | 0.00% | 479 | 0 | 0.00% | 10/29/20 |
| R325125 | 7 | 24 | 0 | 0.00% | 168 | 0 | 0.00% | 10/29/20 |
| R343828 | 12 | 19 | 2 | 16.67% | 230 | 9 | 3.91% | 10/30/20 |
| R325147 | 14 | 21 | 0 | 0.00% | 287 | 0 | 0.00% | 10/30/20 |
| R339853 | 8 | 23 | 0 | 0.00% | 184 | 0 | 0.00% | 10/30/20 |
| R343003 | 16 | 24 | 0 | 0.00% | 378 | 0 | 0.00% | 10/30/20 |

| | | | | | | | | |
|---|---|---|---|---|---|---|---|---|
| R324833 | 55 | 20 | 0 | 0.00% | 1096 | 0 | 0.00% | 10/30/20 |
| R334362 | 1 | 13 | 0 | 0.00% | 13 | 0 | 0.00% | 10/30/20 |
| R325136 | 5 | 22 | 0 | 0.00% | 110 | 0 | 0.00% | 10/30/20 |
| R342986 | 6 | 21 | 0 | 0.00% | 125 | 0 | 0.00% | 10/30/20 |
| R324727 | 1 | 26 | 0 | 0.00% | 26 | 0 | 0.00% | 10/30/20 |
| R325137 | 32 | 24 | 0 | 0.00% | 766 | 0 | 0.00% | 10/30/20 |
| R325529 | 3 | 18 | 0 | 0.00% | 55 | 0 | 0.00% | 10/30/20 |
| R341834 | 4 | 24 | 0 | 0.00% | 96 | 0 | 0.00% | 10/30/20 |
| R343828 | 7 | 17 | 0 | 0.00% | 122 | 0 | 0.00% | 10/31/20 |
| R325127 | 22 | 23 | 1 | 4.55% | 507 | 4 | 0.79% | 11/2/20 |
| R325134 | 5 | 22 | 0 | 0.00% | 111 | 0 | 0.00% | 11/2/20 |
| R325249 | 11 | 24 | 0 | 0.00% | 264 | 0 | 0.00% | 11/2/20 |
| R324727 | 9 | 24 | 0 | 0.00% | 218 | 0 | 0.00% | 11/2/20 |
| R324851 | 43 | 24 | 0 | 0.00% | 1033 | 0 | 0.00% | 11/2/20 |
| R325125 | 43 | 24 | 3 | 6.98% | 1032 | 3 | 0.29% | 11/3/20 |
| R425408 | 51 | 24 | 2 | 3.92% | 1212 | 1 | 0.08% | 11/3/20 |
| R325241 | 51 | 20 | 1 | 1.96% | 1032 | 1 | 0.10% | 11/3/20 |
| R341827 | 5 | 18 | 0 | 0.00% | 90 | 0 | 0.00% | 11/3/20 |
| R341823 | 1 | 11 | 0 | 0.00% | 11 | 0 | 0.00% | 11/3/20 |
| R325134 | 9 | 24 | 0 | 0.00% | 215 | 0 | 0.00% | 11/3/20 |
| R325127 | 12 | 22 | 0 | 0.00% | 258 | 0 | 0.00% | 11/3/20 |
| R325139 | 17 | 20 | 0 | 0.00% | 347 | 0 | 0.00% | 11/3/20 |
| R325138 | 34 | 23 | 0 | 0.00% | 796 | 0 | 0.00% | 11/3/20 |
| R325529 | 4 | 24 | 0 | 0.00% | 96 | 0 | 0.00% | 11/3/20 |
| R341057 | 28 | 18 | 0 | 0.00% | 513 | 0 | 0.00% | 11/3/20 |
| R334327 | 5 | 23 | 4 | 80.00% | 115 | 6 | 5.22% | 11/4/20 |
| R334362 | 36 | 23 | 2 | 5.56% | 820 | 2 | 0.24% | 11/4/20 |
| R425408 | 20 | 24 | 1 | 5.00% | 480 | 2 | 0.42% | 11/4/20 |
| R341827 | 7 | 18 | 0 | 0.00% | 123 | 0 | 0.00% | 11/4/20 |
| R325147 | 13 | 23 | 0 | 0.00% | 304 | 0 | 0.00% | 11/4/20 |
| R426326 | 13 | 24 | 0 | 0.00% | 317 | 0 | 0.00% | 11/4/20 |
| R339853 | 17 | 24 | 0 | 0.00% | 400 | 0 | 0.00% | 11/4/20 |
| R325249 | 9 | 22 | 0 | 0.00% | 195 | 0 | 0.00% | 11/4/20 |
| R334335 | 4 | 19 | 0 | 0.00% | 75 | 0 | 0.00% | 11/4/20 |
| R325136 | 3 | 24 | 0 | 0.00% | 71 | 0 | 0.00% | 11/4/20 |
| R426349 | 19 | 24 | 0 | 0.00% | 449 | 0 | 0.00% | 11/4/20 |
| R325139 | 14 | 24 | 0 | 0.00% | 332 | 0 | 0.00% | 11/4/20 |
| R334342 | 4 | 24 | 0 | 0.00% | 96 | 0 | 0.00% | 11/4/20 |
| R343002 | 13 | 21 | 0 | 0.00% | 272 | 0 | 0.00% | 11/4/20 |
| R325125 | 29 | 24 | 0 | 0.00% | 696 | 0 | 0.00% | 11/4/20 |
| R334324 | 3 | 19 | 0 | 0.00% | 57 | 0 | 0.00% | 11/4/20 |
| R339851 | 30 | 23 | 4 | 13.33% | 690 | 4 | 0.58% | 11/5/20 |
| R325305 | 59 | 24 | 3 | 5.08% | 1406 | 5 | 0.36% | 11/5/20 |
| R334327 | 3 | 19 | 2 | 66.67% | 57 | 2 | 3.51% | 11/5/20 |
| R325129 | 7 | 22 | 1 | 14.29% | 156 | 1 | 0.64% | 11/5/20 |
| R324833 | 55 | 20 | 1 | 1.82% | 1095 | 1 | 0.09% | 11/5/20 |
| R341827 | 5 | 20 | 0 | 0.00% | 99 | 0 | 0.00% | 11/5/20 |
| R325155 | 6 | 24 | 0 | 0.00% | 144 | 0 | 0.00% | 11/5/20 |
| R325147 | 18 | 23 | 0 | 0.00% | 412 | 0 | 0.00% | 11/5/20 |
| R326820 | 7 | 24 | 0 | 0.00% | 166 | 0 | 0.00% | 11/5/20 |
| R325134 | 10 | 24 | 0 | 0.00% | 240 | 0 | 0.00% | 11/5/20 |
| R339422 | 4 | 22 | 0 | 0.00% | 86 | 0 | 0.00% | 11/5/20 |
| R325128 | 16 | 23 | 0 | 0.00% | 366 | 0 | 0.00% | 11/5/20 |
| R325755 | 7 | 22 | 0 | 0.00% | 156 | 0 | 0.00% | 11/5/20 |
| R342986 | 2 | 24 | 0 | 0.00% | 48 | 0 | 0.00% | 11/5/20 |
| R325139 | 9 | 23 | 0 | 0.00% | 205 | 0 | 0.00% | 11/5/20 |
| R324836 | 50 | 23 | 0 | 0.00% | 1128 | 0 | 0.00% | 11/5/20 |
| R325145 | 3 | 24 | 0 | 0.00% | 71 | 0 | 0.00% | 11/5/20 |

| | | | | | | | | |
|---|---|---|---|---|---|---|---|---|
| R325151 | 21 | 23 | 0 | 0.00% | 479 | 0 | 0.00% | 11/5/20 |
| R341834 | 2 | 24 | 0 | 0.00% | 47 | 0 | 0.00% | 11/5/20 |
| R325125 | 8 | 23 | 0 | 0.00% | 185 | 0 | 0.00% | 11/5/20 |
| R341826 | 18 | 21 | 0 | 0.00% | 380 | 0 | 0.00% | 11/5/20 |
| R425408 | 38 | 24 | 0 | 0.00% | 918 | 0 | 0.00% | 11/5/20 |
| R342986 | 6 | 22 | 1 | 16.67% | 130 | 1 | 0.77% | 11/6/20 |
| R325147 | 13 | 22 | 0 | 0.00% | 286 | 0 | 0.00% | 11/6/20 |
| R326820 | 7 | 20 | 0 | 0.00% | 137 | 0 | 0.00% | 11/6/20 |
| R426326 | 15 | 23 | 0 | 0.00% | 338 | 0 | 0.00% | 11/6/20 |
| R325249 | 8 | 24 | 0 | 0.00% | 190 | 0 | 0.00% | 11/6/20 |
| R343003 | 19 | 22 | 0 | 0.00% | 412 | 0 | 0.00% | 11/6/20 |
| R334362 | 1 | 8 | 0 | 0.00% | 8 | 0 | 0.00% | 11/6/20 |
| R325136 | 2 | 24 | 0 | 0.00% | 48 | 0 | 0.00% | 11/6/20 |
| R325544 | 2 | 13 | 0 | 0.00% | 26 | 0 | 0.00% | 11/6/20 |
| R325137 | 10 | 24 | 0 | 0.00% | 240 | 0 | 0.00% | 11/6/20 |
| R325570 | 1 | 18 | 0 | 0.00% | 18 | 0 | 0.00% | 11/6/20 |
| R325529 | 12 | 23 | 0 | 0.00% | 276 | 0 | 0.00% | 11/6/20 |
| R324851 | 4 | 22 | 0 | 0.00% | 88 | 0 | 0.00% | 11/6/20 |
| R326854 | 1 | 26 | 0 | 0.00% | 26 | 0 | 0.00% | 11/6/20 |
| R325125 | 6 | 23 | 0 | 0.00% | 140 | 0 | 0.00% | 11/6/20 |
| R325137 | 16 | 23 | 0 | 0.00% | 366 | 0 | 0.00% | 11/7/20 |
| R334327 | 2 | 22 | 0 | 0.00% | 44 | 0 | 0.00% | 11/7/20 |
| R325127 | 27 | 22 | 4 | 14.81% | 592 | 4 | 0.68% | 11/9/20 |
| R325129 | 2 | 14 | 2 | 100.00% | 28 | 2 | 7.14% | 11/9/20 |
| R325249 | 7 | 22 | 0 | 0.00% | 157 | 0 | 0.00% | 11/9/20 |
| R342986 | 6 | 21 | 0 | 0.00% | 128 | 0 | 0.00% | 11/9/20 |
| R325148 | 6 | 23 | 0 | 0.00% | 137 | 0 | 0.00% | 11/9/20 |
| R334327 | 4 | 24 | 0 | 0.00% | 96 | 0 | 0.00% | 11/9/20 |
| R425408 | 64 | 24 | 10 | 15.63% | 1533 | 11 | 0.72% | 11/10/20 |
| R339851 | 45 | 23 | 4 | 8.89% | 1055 | 4 | 0.38% | 11/10/20 |
| R325241 | 58 | 17 | 4 | 6.90% | 989 | 1 | 0.10% | 11/10/20 |
| R325125 | 38 | 24 | 1 | 2.63% | 912 | 2 | 0.22% | 11/10/20 |
| R341849 | 1 | 24 | 0 | 0.00% | 24 | 0 | 0.00% | 11/10/20 |
| R341827 | 6 | 18 | 0 | 0.00% | 105 | 0 | 0.00% | 11/10/20 |
| R341823 | 1 | 10 | 0 | 0.00% | 10 | 0 | 0.00% | 11/10/20 |
| R325134 | 5 | 24 | 0 | 0.00% | 120 | 0 | 0.00% | 11/10/20 |
| R343828 | 7 | 24 | 0 | 0.00% | 165 | 0 | 0.00% | 11/10/20 |
| R324727 | 8 | 22 | 0 | 0.00% | 178 | 0 | 0.00% | 11/10/20 |
| R325127 | 17 | 22 | 0 | 0.00% | 371 | 0 | 0.00% | 11/10/20 |
| R426349 | 17 | 23 | 0 | 0.00% | 398 | 0 | 0.00% | 11/10/20 |
| R325138 | 33 | 24 | 0 | 0.00% | 778 | 0 | 0.00% | 11/10/20 |
| R325529 | 4 | 24 | 0 | 0.00% | 96 | 0 | 0.00% | 11/10/20 |
| R341057 | 27 | 20 | 0 | 0.00% | 528 | 0 | 0.00% | 11/10/20 |
| R325129 | 5 | 23 | 2 | 40.00% | 113 | 2 | 1.77% | 11/11/20 |
| R339853 | 12 | 24 | 2 | 16.67% | 292 | 2 | 0.68% | 11/11/20 |
| R325249 | 13 | 23 | 2 | 15.38% | 295 | 2 | 0.68% | 11/11/20 |
| R341826 | 18 | 23 | 2 | 11.11% | 414 | 2 | 0.48% | 11/11/20 |
| R334362 | 36 | 22 | 2 | 5.56% | 803 | 2 | 0.25% | 11/11/20 |
| R325125 | 36 | 24 | 2 | 5.56% | 864 | 3 | 0.35% | 11/11/20 |
| R425408 | 34 | 23 | 1 | 2.94% | 798 | 1 | 0.13% | 11/11/20 |
| R341831 | 3 | 23 | 0 | 0.00% | 70 | 0 | 0.00% | 11/11/20 |
| R325147 | 16 | 21 | 0 | 0.00% | 339 | 0 | 0.00% | 11/11/20 |
| R339422 | 4 | 23 | 0 | 0.00% | 90 | 0 | 0.00% | 11/11/20 |
| R426326 | 14 | 23 | 0 | 0.00% | 327 | 0 | 0.00% | 11/11/20 |
| R341850 | 13 | 21 | 0 | 0.00% | 272 | 0 | 0.00% | 11/11/20 |
| R341840 | 20 | 21 | 0 | 0.00% | 416 | 0 | 0.00% | 11/11/20 |
| R325136 | 2 | 24 | 0 | 0.00% | 48 | 0 | 0.00% | 11/11/20 |
| R334342 | 4 | 23 | 0 | 0.00% | 90 | 0 | 0.00% | 11/11/20 |

| | | | | | | | | |
|---|---|---|---|---|---|---|---|---|
| R325529 | 5 | 22 | 0 | 0.00% | 108 | 0 | 0.00% | 11/11/20 |
| R343002 | 12 | 23 | 0 | 0.00% | 271 | 0 | 0.00% | 11/11/20 |
| R324851 | 6 | 23 | 0 | 0.00% | 135 | 0 | 0.00% | 11/11/20 |
| R326854 | 1 | 22 | 0 | 0.00% | 22 | 0 | 0.00% | 11/11/20 |
| R341867 | 3 | 20 | 0 | 0.00% | 61 | 0 | 0.00% | 11/11/20 |
| R334324 | 3 | 19 | 0 | 0.00% | 57 | 0 | 0.00% | 11/11/20 |
| R325139 | 16 | 22 | 5 | 31.25% | 349 | 7 | 2.01% | 11/12/20 |
| R325139 | 30 | 22 | 5 | 31.25% | 674 | 7 | 1.04% | 11/12/20 |
| R325139 | 30 | 12 | 5 | 16.67% | 349 | 7 | 2.01% | 11/12/20 |
| R325139 | 30 | 22 | 5 | 16.67% | 674 | 7 | 1.04% | 11/12/20 |
| R341834 | 2 | 24 | 2 | 100.00% | 48 | 2 | 4.17% | 11/12/20 |
| R324836 | 56 | 18 | 2 | 3.57% | 1011 | 2 | 0.20% | 11/12/20 |
| R325125 | 7 | 24 | 1 | 14.29% | 168 | 1 | 0.60% | 11/12/20 |
| R325155 | 7 | 24 | 0 | 0.00% | 168 | 0 | 0.00% | 11/12/20 |
| R325147 | 18 | 22 | 0 | 0.00% | 391 | 0 | 0.00% | 11/12/20 |
| R325554 | 4 | 23 | 0 | 0.00% | 91 | 0 | 0.00% | 11/12/20 |
| R325134 | 11 | 24 | 0 | 0.00% | 264 | 0 | 0.00% | 11/12/20 |
| R325128 | 16 | 23 | 0 | 0.00% | 363 | 0 | 0.00% | 11/12/20 |
| R325755 | 6 | 26 | 0 | 0.00% | 153 | 0 | 0.00% | 11/12/20 |
| R342986 | 4 | 24 | 0 | 0.00% | 96 | 0 | 0.00% | 11/12/20 |
| R334327 | 2 | 24 | 0 | 0.00% | 48 | 0 | 0.00% | 11/12/20 |
| R325145 | 2 | 24 | 0 | 0.00% | 47 | 0 | 0.00% | 11/12/20 |
| R325151 | 20 | 24 | 0 | 0.00% | 477 | 0 | 0.00% | 11/12/20 |
| R325137 | 34 | 24 | 6 | 17.65% | 811 | 6 | 0.74% | 11/13/20 |
| R325305 | 61 | 23 | 6 | 9.84% | 1387 | 7 | 0.50% | 11/13/20 |
| R324833 | 55 | 20 | 4 | 7.27% | 1088 | 5 | 0.46% | 11/13/20 |
| R426326 | 14 | 23 | 3 | 21.43% | 326 | 4 | 1.23% | 11/13/20 |
| R343003 | 17 | 23 | 2 | 11.76% | 391 | 2 | 0.51% | 11/13/20 |
| R325147 | 12 | 23 | 0 | 0.00% | 273 | 0 | 0.00% | 11/13/20 |
| R325249 | 8 | 23 | 0 | 0.00% | 182 | 0 | 0.00% | 11/13/20 |
| R334362 | 1 | 9 | 0 | 0.00% | 9 | 0 | 0.00% | 11/13/20 |
| R325136 | 3 | 24 | 0 | 0.00% | 72 | 0 | 0.00% | 11/13/20 |
| R342986 | 3 | 23 | 0 | 0.00% | 69 | 0 | 0.00% | 11/13/20 |
| R325544 | 2 | 14 | 0 | 0.00% | 28 | 0 | 0.00% | 11/13/20 |
| R325529 | 8 | 23 | 0 | 0.00% | 183 | 0 | 0.00% | 11/13/20 |
| R325125 | 8 | 24 | 0 | 0.00% | 192 | 0 | 0.00% | 11/13/20 |
| R325127 | 28 | 22 | 1 | 3.57% | 608 | 1 | 0.16% | 11/16/20 |
| R325249 | 8 | 24 | 0 | 0.00% | 193 | 0 | 0.00% | 11/16/20 |
| R324727 | 8 | 24 | 0 | 0.00% | 190 | 0 | 0.00% | 11/16/20 |
| R325529 | 4 | 24 | 0 | 0.00% | 95 | 0 | 0.00% | 11/16/20 |
| R324851 | 19 | 24 | 0 | 0.00% | 450 | 0 | 0.00% | 11/16/20 |
| R325125 | 5 | 22 | 0 | 0.00% | 111 | 0 | 0.00% | 11/16/20 |
| R325125 | 38 | 24 | 2 | 5.26% | 903 | 2 | 0.22% | 11/17/20 |
| R325241 | 58 | 17 | 2 | 3.45% | 981 | 2 | 0.20% | 11/17/20 |
| R426326 | 1 | 2 | 1 | 100.00% | 2 | 2 | 100.00% | 11/17/20 |
| R325139 | 18 | 23 | 1 | 5.56% | 415 | 1 | 0.24% | 11/17/20 |
| R425408 | 41 | 23 | 1 | 2.44% | 963 | 2 | 0.21% | 11/17/20 |
| R341827 | 5 | 17 | 0 | 0.00% | 86 | 0 | 0.00% | 11/17/20 |
| R341823 | 2 | 13 | 0 | 0.00% | 26 | 0 | 0.00% | 11/17/20 |
| R325134 | 11 | 23 | 0 | 0.00% | 258 | 0 | 0.00% | 11/17/20 |
| R339422 | 4 | 22 | 0 | 0.00% | 87 | 0 | 0.00% | 11/17/20 |
| R325755 | 3 | 23 | 0 | 0.00% | 69 | 0 | 0.00% | 11/17/20 |
| R325136 | 3 | 24 | 0 | 0.00% | 72 | 0 | 0.00% | 11/17/20 |
| R325127 | 18 | 21 | 0 | 0.00% | 383 | 0 | 0.00% | 11/17/20 |
| R325129 | 6 | 23 | 0 | 0.00% | 140 | 0 | 0.00% | 11/17/20 |
| R426349 | 19 | 23 | 0 | 0.00% | 435 | 0 | 0.00% | 11/17/20 |
| R325138 | 28 | 24 | 0 | 0.00% | 670 | 0 | 0.00% | 11/17/20 |
| R339851 | 23 | 23 | 0 | 0.00% | 540 | 0 | 0.00% | 11/17/20 |

| | | | | | | | | |
|---|---|---|---|---|---|---|---|---|
| R334327 | 2 | 23 | 0 | 0.00% | 46 | 0 | 0.00% | 11/17/20 |
| R325129 | 5 | 20 | 2 | 40.00% | 101 | 2 | 1.98% | 11/18/20 |
| R426326 | 8 | 22 | 1 | 12.50% | 177 | 0 | 0.00% | 11/18/20 |
| R341827 | 10 | 20 | 1 | 10.00% | 195 | 1 | 0.51% | 11/18/20 |
| R341850 | 13 | 21 | 1 | 7.69% | 272 | 1 | 0.37% | 11/18/20 |
| R339853 | 17 | 22 | 1 | 5.88% | 379 | 1 | 0.26% | 11/18/20 |
| R325249 | 17 | 23 | 1 | 5.88% | 386 | 1 | 0.26% | 11/18/20 |
| R325125 | 37 | 24 | 1 | 2.70% | 888 | 1 | 0.11% | 11/18/20 |
| R325147 | 17 | 21 | 0 | 0.00% | 361 | 0 | 0.00% | 11/18/20 |
| R343828 | 8 | 20 | 0 | 0.00% | 161 | 0 | 0.00% | 11/18/20 |
| R339308 | 2 | 22 | 0 | 0.00% | 44 | 0 | 0.00% | 11/18/20 |
| R341840 | 20 | 21 | 0 | 0.00% | 429 | 0 | 0.00% | 11/18/20 |
| R334362 | 36 | 22 | 0 | 0.00% | 800 | 0 | 0.00% | 11/18/20 |
| R334342 | 4 | 24 | 0 | 0.00% | 96 | 0 | 0.00% | 11/18/20 |
| R325148 | 6 | 23 | 0 | 0.00% | 138 | 0 | 0.00% | 11/18/20 |
| R325529 | 9 | 24 | 0 | 0.00% | 216 | 0 | 0.00% | 11/18/20 |
| R325145 | 3 | 21 | 0 | 0.00% | 63 | 0 | 0.00% | 11/18/20 |
| R343002 | 12 | 22 | 0 | 0.00% | 264 | 0 | 0.00% | 11/18/20 |
| R341825 | 5 | 16 | 0 | 0.00% | 79 | 0 | 0.00% | 11/18/20 |
| R341834 | 2 | 20 | 0 | 0.00% | 39 | 0 | 0.00% | 11/18/20 |
| R341057 | 21 | 23 | 0 | 0.00% | 481 | 0 | 0.00% | 11/18/20 |
| R324851 | 45 | 24 | 0 | 0.00% | 1058 | 0 | 0.00% | 11/18/20 |
| R341867 | 3 | 23 | 0 | 0.00% | 68 | 0 | 0.00% | 11/18/20 |
| R425408 | 17 | 23 | 0 | 0.00% | 390 | 0 | 0.00% | 11/18/20 |
| R334324 | 3 | 18 | 0 | 0.00% | 54 | 0 | 0.00% | 11/18/20 |
| R324836 | 45 | 23 | 2 | 4.44% | 1013 | 1 | 0.10% | 11/19/20 |
| R334335 | 3 | 23 | 1 | 33.33% | 70 | 1 | 1.43% | 11/19/20 |
| R325305 | 40 | 22 | 1 | 2.50% | 874 | 3 | 0.34% | 11/19/20 |
| R341827 | 7 | 19 | 0 | 0.00% | 131 | 0 | 0.00% | 11/19/20 |
| R325155 | 13 | 23 | 0 | 0.00% | 300 | 0 | 0.00% | 11/19/20 |
| R325147 | 16 | 22 | 0 | 0.00% | 357 | 0 | 0.00% | 11/19/20 |
| R325134 | 11 | 22 | 0 | 0.00% | 242 | 0 | 0.00% | 11/19/20 |
| R325128 | 16 | 23 | 0 | 0.00% | 367 | 0 | 0.00% | 11/19/20 |
| R325755 | 5 | 22 | 0 | 0.00% | 111 | 0 | 0.00% | 11/19/20 |
| R325136 | 2 | 20 | 0 | 0.00% | 39 | 0 | 0.00% | 11/19/20 |
| R342986 | 8 | 22 | 0 | 0.00% | 174 | 0 | 0.00% | 11/19/20 |
| R339301 | 7 | 19 | 0 | 0.00% | 132 | 0 | 0.00% | 11/19/20 |
| R325125 | 2 | 24 | 0 | 0.00% | 48 | 0 | 0.00% | 11/19/20 |
| R325137 | 33 | 24 | 4 | 12.12% | 791 | 5 | 0.63% | 11/20/20 |
| R426326 | 7 | 21 | 1 | 14.29% | 145 | 2 | 1.38% | 11/20/20 |
| R325249 | 11 | 22 | 1 | 9.09% | 241 | 1 | 0.41% | 11/20/20 |
| R325125 | 11 | 24 | 1 | 9.09% | 260 | 1 | 0.38% | 11/20/20 |
| R325147 | 13 | 23 | 1 | 7.69% | 298 | 1 | 0.34% | 11/20/20 |
| R341866 | 29 | 24 | 1 | 3.45% | 690 | 2 | 0.29% | 11/20/20 |
| R341858 | 31 | 24 | 1 | 3.23% | 743 | 1 | 0.13% | 11/20/20 |
| R343003 | 17 | 23 | 0 | 0.00% | 386 | 0 | 0.00% | 11/20/20 |
| R324833 | 55 | 19 | 0 | 0.00% | 1068 | 0 | 0.00% | 11/20/20 |
| R343386 | 2 | 24 | 0 | 0.00% | 48 | 0 | 0.00% | 11/20/20 |
| R325544 | 2 | 14 | 0 | 0.00% | 28 | 0 | 0.00% | 11/20/20 |
| R326854 | 1 | 22 | 0 | 0.00% | 22 | 0 | 0.00% | 11/20/20 |
| R339851 | 21 | 24 | 1 | 4.76% | 497 | 1 | 0.20% | 11/23/20 |
| R325249 | 10 | 23 | 0 | 0.00% | 225 | 0 | 0.00% | 11/23/20 |
| R334324 | 3 | 17 | 0 | 0.00% | 50 | 0 | 0.00% | 11/23/20 |
| R325134 | 5 | 22 | 1 | 20.00% | 108 | 1 | 0.93% | 11/24/20 |
| R324855 | 2 | 15 | 0 | 0.00% | 29 | 0 | 0.00% | 11/24/20 |
| R325129 | 7 | 22 | 0 | 0.00% | 157 | 0 | 0.00% | 11/24/20 |
| R339483 | 8 | 20 | 0 | 0.00% | 156 | 0 | 0.00% | 11/24/20 |
| R426349 | 12 | 24 | 0 | 0.00% | 282 | 0 | 0.00% | 11/24/20 |

| | | | | | | | | |
|---|---|---|---|---|---|---|---|---|
| R325125 | 16 | 24 | 0 | 0.00% | 377 | 0 | 0.00% | 11/24/20 |
| R425408 | 40 | 24 | 0 | 0.00% | 946 | 0 | 0.00% | 11/24/20 |
| R325249 | 3 | 22 | 1 | 33.33% | 66 | 1 | 1.52% | 11/25/20 |
| R425408 | 18 | 23 | 1 | 5.56% | 416 | 1 | 0.24% | 11/25/20 |
| R342167 | 1 | 19 | 0 | 0.00% | 19 | 0 | 0.00% | 11/25/20 |
| R334335 | 3 | 19 | 0 | 0.00% | 58 | 0 | 0.00% | 11/25/20 |
| R339308 | 2 | 24 | 0 | 0.00% | 48 | 0 | 0.00% | 11/25/20 |
| R325137 | 4 | 22 | 0 | 0.00% | 87 | 0 | 0.00% | 11/25/20 |
| R339492 | 4 | 16 | 0 | 0.00% | 62 | 0 | 0.00% | 11/25/20 |
| R324833 | 7 | 20 | 0 | 0.00% | 140 | 0 | 0.00% | 11/27/20 |
| R426787 | 35 | 22 | 4 | 11.43% | 769 | 4 | 0.52% | 11/30/20 |
| R325241 | 42 | 23 | 3 | 7.14% | 968 | 3 | 0.31% | 11/30/20 |
| R425408 | 52 | 23 | 2 | 3.85% | 1196 | 4 | 0.33% | 11/30/20 |
| R426665 | 8 | 22 | 1 | 12.50% | 174 | 1 | 0.57% | 11/30/20 |
| R325249 | 14 | 22 | 1 | 7.14% | 308 | 1 | 0.32% | 11/30/20 |
| R325127 | 44 | 23 | 1 | 2.27% | 1029 | 1 | 0.10% | 11/30/20 |
| R326820 | 14 | 23 | 0 | 0.00% | 325 | 0 | 0.00% | 11/30/20 |
| R341858 | 30 | 24 | 0 | 0.00% | 720 | 0 | 0.00% | 11/30/20 |
| R334342 | 18 | 24 | 0 | 0.00% | 427 | 0 | 0.00% | 11/30/20 |
| R341057 | 15 | 22 | 0 | 0.00% | 329 | 0 | 0.00% | 11/30/20 |
| R325125 | 92 | 24 | 14 | 15.22% | 2202 | 19 | 0.86% | 12/1/20 |
| R343003 | 11 | 23 | 2 | 18.18% | 251 | 2 | 0.80% | 12/1/20 |
| R325127 | 2 | 15 | 1 | 50.00% | 30 | 1 | 3.33% | 12/1/20 |
| R325129 | 6 | 23 | 1 | 16.67% | 135 | 1 | 0.74% | 12/1/20 |
| R383214 | 21 | 22 | 1 | 4.76% | 468 | 1 | 0.21% | 12/1/20 |
| R341827 | 36 | 20 | 1 | 2.78% | 711 | 1 | 0.14% | 12/1/20 |
| R325147 | 4 | 21 | 0 | 0.00% | 83 | 0 | 0.00% | 12/1/20 |
| R383220 | 5 | 13 | 0 | 0.00% | 67 | 0 | 0.00% | 12/1/20 |
| R325134 | 9 | 23 | 0 | 0.00% | 207 | 0 | 0.00% | 12/1/20 |
| R339422 | 4 | 21 | 0 | 0.00% | 83 | 0 | 0.00% | 12/1/20 |
| R383226 | 3 | 12 | 0 | 0.00% | 35 | 0 | 0.00% | 12/1/20 |
| R334327 | 4 | 21 | 0 | 0.00% | 84 | 0 | 0.00% | 12/1/20 |
| R341057 | 11 | 21 | 0 | 0.00% | 229 | 0 | 0.00% | 12/1/20 |
| R382253 | 30 | 24 | 3 | 10.00% | 710 | 2 | 0.28% | 12/2/20 |
| R341866 | 28 | 24 | 2 | 7.14% | 664 | 2 | 0.30% | 12/2/20 |
| R325129 | 5 | 21 | 1 | 20.00% | 103 | 2 | 1.94% | 12/2/20 |
| R383622 | 6 | 15 | 0 | 0.00% | 89 | 0 | 0.00% | 12/2/20 |
| R341827 | 15 | 19 | 0 | 0.00% | 289 | 0 | 0.00% | 12/2/20 |
| R325147 | 12 | 22 | 0 | 0.00% | 265 | 0 | 0.00% | 12/2/20 |
| R383220 | 5 | 16 | 0 | 0.00% | 80 | 0 | 0.00% | 12/2/20 |
| R325554 | 16 | 24 | 0 | 0.00% | 384 | 0 | 0.00% | 12/2/20 |
| R325249 | 16 | 20 | 0 | 0.00% | 326 | 0 | 0.00% | 12/2/20 |
| R324855 | 1 | 29 | 0 | 0.00% | 29 | 0 | 0.00% | 12/2/20 |
| R341850 | 13 | 19 | 0 | 0.00% | 241 | 0 | 0.00% | 12/2/20 |
| R334335 | 3 | 24 | 0 | 0.00% | 71 | 0 | 0.00% | 12/2/20 |
| R341840 | 5 | 24 | 0 | 0.00% | 118 | 0 | 0.00% | 12/2/20 |
| R325139 | 34 | 24 | 0 | 0.00% | 829 | 0 | 0.00% | 12/2/20 |
| R382442 | 30 | 24 | 0 | 0.00% | 710 | 0 | 0.00% | 12/2/20 |
| R383214 | 1 | 7 | 0 | 0.00% | 7 | 0 | 0.00% | 12/2/20 |
| R343002 | 12 | 23 | 0 | 0.00% | 271 | 0 | 0.00% | 12/2/20 |
| R325125 | 2 | 20 | 0 | 0.00% | 40 | 0 | 0.00% | 12/2/20 |
| R425408 | 40 | 23 | 0 | 0.00% | 934 | 0 | 0.00% | 12/2/20 |
| R334324 | 3 | 18 | 0 | 0.00% | 53 | 0 | 0.00% | 12/2/20 |
| R324836 | 56 | 19 | 4 | 7.14% | 1069 | 4 | 0.37% | 12/3/20 |
| R382442 | 15 | 22 | 3 | 20.00% | 335 | 4 | 1.19% | 12/3/20 |
| R325305 | 58 | 23 | 3 | 5.17% | 1347 | 2 | 0.15% | 12/3/20 |
| R325137 | 59 | 24 | 3 | 5.08% | 1401 | 3 | 0.21% | 12/3/20 |
| R424259 | 5 | 22 | 2 | 40.00% | 110 | 3 | 2.73% | 12/3/20 |

| | | | | | | | | |
|---|---|---|---|---|---|---|---|---|
| R325554 | 17 | 23 | 2 | 11.76% | 399 | 2 | 0.50% | 12/3/20 |
| R342986 | 3 | 10 | 1 | 33.33% | 31 | 1 | 3.23% | 12/3/20 |
| R339478 | 23 | 24 | 1 | 4.35% | 548 | 1 | 0.18% | 12/3/20 |
| R325127 | 25 | 23 | 1 | 4.00% | 573 | 1 | 0.17% | 12/3/20 |
| R341057 | 26 | 22 | 1 | 3.85% | 561 | 1 | 0.18% | 12/3/20 |
| R341827 | 7 | 18 | 0 | 0.00% | 125 | 0 | 0.00% | 12/3/20 |
| R325147 | 4 | 20 | 0 | 0.00% | 79 | 0 | 0.00% | 12/3/20 |
| R326820 | 1 | 16 | 0 | 0.00% | 16 | 0 | 0.00% | 12/3/20 |
| R325134 | 10 | 24 | 0 | 0.00% | 239 | 0 | 0.00% | 12/3/20 |
| R324855 | 9 | 24 | 0 | 0.00% | 214 | 0 | 0.00% | 12/3/20 |
| R325128 | 17 | 24 | 0 | 0.00% | 400 | 0 | 0.00% | 12/3/20 |
| R423648 | 12 | 23 | 0 | 0.00% | 281 | 0 | 0.00% | 12/3/20 |
| R325129 | 1 | 19 | 0 | 0.00% | 19 | 0 | 0.00% | 12/3/20 |
| R339483 | 8 | 19 | 0 | 0.00% | 155 | 0 | 0.00% | 12/3/20 |
| R423627 | 8 | 24 | 0 | 0.00% | 188 | 0 | 0.00% | 12/3/20 |
| R423633 | 2 | 24 | 0 | 0.00% | 48 | 0 | 0.00% | 12/3/20 |
| R342151 | 5 | 19 | 0 | 0.00% | 95 | 0 | 0.00% | 12/3/20 |
| R425408 | 16 | 23 | 0 | 0.00% | 372 | 0 | 0.00% | 12/3/20 |
| R325125 | 21 | 24 | 6 | 28.57% | 502 | 1 | 0.20% | 12/4/20 |
| R382442 | 24 | 23 | 3 | 12.50% | 547 | 5 | 0.91% | 12/4/20 |
| R340789 | 7 | 17 | 1 | 14.29% | 122 | 1 | 0.82% | 12/4/20 |
| R424384 | 9 | 22 | 1 | 11.11% | 201 | 1 | 0.50% | 12/4/20 |
| R325529 | 18 | 22 | 1 | 5.56% | 403 | 1 | 0.25% | 12/4/20 |
| R325147 | 22 | 21 | 1 | 4.55% | 451 | 1 | 0.22% | 12/4/20 |
| R341823 | 2 | 16 | 0 | 0.00% | 32 | 0 | 0.00% | 12/4/20 |
| R325134 | 2 | 17 | 0 | 0.00% | 33 | 0 | 0.00% | 12/4/20 |
| R342167 | 1 | 22 | 0 | 0.00% | 22 | 0 | 0.00% | 12/4/20 |
| R325249 | 14 | 22 | 0 | 0.00% | 310 | 0 | 0.00% | 12/4/20 |
| R343003 | 10 | 24 | 0 | 0.00% | 240 | 0 | 0.00% | 12/4/20 |
| R324833 | 49 | 21 | 0 | 0.00% | 1045 | 0 | 0.00% | 12/4/20 |
| R325755 | 6 | 21 | 0 | 0.00% | 126 | 0 | 0.00% | 12/4/20 |
| R342986 | 4 | 19 | 0 | 0.00% | 77 | 0 | 0.00% | 12/4/20 |
| R325127 | 15 | 23 | 0 | 0.00% | 349 | 0 | 0.00% | 12/4/20 |
| R341858 | 31 | 24 | 0 | 0.00% | 729 | 0 | 0.00% | 12/4/20 |
| R423627 | 7 | 20 | 0 | 0.00% | 137 | 0 | 0.00% | 12/4/20 |
| R334342 | 4 | 24 | 0 | 0.00% | 94 | 0 | 0.00% | 12/4/20 |
| R339485 | 4 | 21 | 0 | 0.00% | 84 | 0 | 0.00% | 12/4/20 |
| R424356 | 4 | 24 | 0 | 0.00% | 95 | 0 | 0.00% | 12/4/20 |
| R324851 | 13 | 24 | 0 | 0.00% | 312 | 0 | 0.00% | 12/4/20 |
| R326854 | 2 | 23 | 0 | 0.00% | 45 | 0 | 0.00% | 12/4/20 |
| R383226 | 2 | 19 | 0 | 0.00% | 38 | 0 | 0.00% | 12/5/20 |
| R325249 | 11 | 23 | 1 | 9.09% | 254 | 1 | 0.39% | 12/7/20 |
| R424410 | 10 | 21 | 0 | 0.00% | 211 | 0 | 0.00% | 12/7/20 |
| R424386 | 7 | 20 | 0 | 0.00% | 140 | 0 | 0.00% | 12/7/20 |
| R339853 | 15 | 24 | 0 | 0.00% | 360 | 0 | 0.00% | 12/7/20 |
| R325127 | 26 | 23 | 0 | 0.00% | 595 | 0 | 0.00% | 12/7/20 |
| R325148 | 6 | 23 | 0 | 0.00% | 140 | 0 | 0.00% | 12/7/20 |
| R325570 | 2 | 18 | 0 | 0.00% | 36 | 0 | 0.00% | 12/7/20 |
| R325125 | 50 | 24 | 4 | 8.00% | 1184 | 5 | 0.42% | 12/8/20 |
| R325129 | 8 | 21 | 1 | 12.50% | 168 | 1 | 0.60% | 12/8/20 |
| R339851 | 17 | 22 | 1 | 5.88% | 374 | 1 | 0.27% | 12/8/20 |
| R325305 | 53 | 24 | 1 | 1.89% | 1261 | 1 | 0.08% | 12/8/20 |
| R325241 | 60 | 16 | 1 | 1.67% | 980 | 2 | 0.20% | 12/8/20 |
| R341827 | 5 | 17 | 0 | 0.00% | 84 | 0 | 0.00% | 12/8/20 |
| R326820 | 8 | 21 | 0 | 0.00% | 165 | 0 | 0.00% | 12/8/20 |
| R325134 | 9 | 24 | 0 | 0.00% | 216 | 0 | 0.00% | 12/8/20 |
| R339422 | 4 | 23 | 0 | 0.00% | 92 | 0 | 0.00% | 12/8/20 |
| R342167 | 1 | 16 | 0 | 0.00% | 16 | 0 | 0.00% | 12/8/20 |

| | | | | | | | | |
|---|---|---|---|---|---|---|---|---|
| R334327 | 3 | 20 | 0 | 0.00% | 59 | 0 | 0.00% | 12/8/20 |
| R339472 | 1 | 21 | 0 | 0.00% | 21 | 0 | 0.00% | 12/8/20 |
| R341057 | 25 | 23 | 0 | 0.00% | 566 | 0 | 0.00% | 12/8/20 |
| R425408 | 46 | 23 | 0 | 0.00% | 1076 | 0 | 0.00% | 12/8/20 |
| R339490 | 1 | 7 | 0 | 0.00% | 7 | 0 | 0.00% | 12/8/20 |
| R325129 | 5 | 19 | 3 | 60.00% | 94 | 3 | 3.19% | 12/9/20 |
| R325127 | 14 | 23 | 2 | 14.29% | 323 | 2 | 0.62% | 12/9/20 |
| R425408 | 55 | 24 | 2 | 3.64% | 1307 | 3 | 0.23% | 12/9/20 |
| R326820 | 5 | 23 | 1 | 20.00% | 115 | 1 | 0.87% | 12/9/20 |
| R325529 | 6 | 23 | 1 | 16.67% | 135 | 1 | 0.74% | 12/9/20 |
| R382442 | 18 | 19 | 1 | 5.56% | 348 | 1 | 0.29% | 12/9/20 |
| R325125 | 20 | 23 | 1 | 5.00% | 456 | 1 | 0.22% | 12/9/20 |
| R341866 | 28 | 23 | 1 | 3.57% | 653 | 1 | 0.15% | 12/9/20 |
| R383622 | 6 | 17 | 0 | 0.00% | 103 | 0 | 0.00% | 12/9/20 |
| R341827 | 9 | 17 | 0 | 0.00% | 156 | 0 | 0.00% | 12/9/20 |
| R325147 | 5 | 16 | 0 | 0.00% | 81 | 0 | 0.00% | 12/9/20 |
| R424410 | 5 | 18 | 0 | 0.00% | 89 | 0 | 0.00% | 12/9/20 |
| R325249 | 15 | 23 | 0 | 0.00% | 352 | 0 | 0.00% | 12/9/20 |
| R341850 | 13 | 20 | 0 | 0.00% | 265 | 0 | 0.00% | 12/9/20 |
| R341840 | 5 | 21 | 0 | 0.00% | 106 | 0 | 0.00% | 12/9/20 |
| R424428 | 6 | 18 | 0 | 0.00% | 108 | 0 | 0.00% | 12/9/20 |
| R339318 | 4 | 20 | 0 | 0.00% | 81 | 0 | 0.00% | 12/9/20 |
| R382276 | 1 | 24 | 0 | 0.00% | 24 | 0 | 0.00% | 12/9/20 |
| R426349 | 10 | 24 | 0 | 0.00% | 237 | 0 | 0.00% | 12/9/20 |
| R325139 | 36 | 22 | 0 | 0.00% | 809 | 0 | 0.00% | 12/9/20 |
| R334342 | 4 | 24 | 0 | 0.00% | 96 | 0 | 0.00% | 12/9/20 |
| R339478 | 12 | 18 | 0 | 0.00% | 210 | 0 | 0.00% | 12/9/20 |
| R422912 | 3 | 22 | 0 | 0.00% | 67 | 0 | 0.00% | 12/9/20 |
| R343002 | 12 | 22 | 0 | 0.00% | 265 | 0 | 0.00% | 12/9/20 |
| R334324 | 3 | 19 | 0 | 0.00% | 58 | 0 | 0.00% | 12/9/20 |
| R324851 | 60 | 24 | 5 | 8.33% | 1442 | 5 | 0.35% | 12/10/20 |
| R325128 | 19 | 24 | 2 | 10.53% | 447 | 2 | 0.45% | 12/10/20 |
| R325305 | 56 | 23 | 2 | 3.57% | 1302 | 1 | 0.08% | 12/10/20 |
| R325129 | 2 | 12 | 1 | 50.00% | 23 | 1 | 4.35% | 12/10/20 |
| R342986 | 6 | 24 | 1 | 16.67% | 145 | 1 | 0.69% | 12/10/20 |
| R324836 | 51 | 20 | 1 | 1.96% | 1029 | 3 | 0.29% | 12/10/20 |
| R341827 | 6 | 16 | 0 | 0.00% | 97 | 0 | 0.00% | 12/10/20 |
| R325147 | 30 | 20 | 0 | 0.00% | 601 | 0 | 0.00% | 12/10/20 |
| R325554 | 13 | 21 | 0 | 0.00% | 267 | 0 | 0.00% | 12/10/20 |
| R325134 | 12 | 23 | 0 | 0.00% | 275 | 0 | 0.00% | 12/10/20 |
| R334335 | 3 | 24 | 0 | 0.00% | 72 | 0 | 0.00% | 12/10/20 |
| R325755 | 8 | 22 | 0 | 0.00% | 174 | 0 | 0.00% | 12/10/20 |
| R339483 | 8 | 20 | 0 | 0.00% | 157 | 0 | 0.00% | 12/10/20 |
| R334327 | 13 | 23 | 0 | 0.00% | 303 | 0 | 0.00% | 12/10/20 |
| R342151 | 5 | 20 | 0 | 0.00% | 98 | 0 | 0.00% | 12/10/20 |
| R325125 | 7 | 24 | 0 | 0.00% | 166 | 0 | 0.00% | 12/10/20 |
| R325125 | 8 | 24 | 3 | 37.50% | 188 | 8 | 4.26% | 12/11/20 |
| R426349 | 13 | 21 | 3 | 23.08% | 270 | 3 | 1.11% | 12/11/20 |
| R382253 | 36 | 20 | 2 | 5.56% | 705 | 2 | 0.28% | 12/11/20 |
| R324851 | 1 | 3 | 1 | 100.00% | 3 | 1 | 33.33% | 12/11/20 |
| R325137 | 11 | 24 | 1 | 9.09% | 264 | 6 | 2.27% | 12/11/20 |
| R382442 | 13 | 24 | 1 | 7.69% | 309 | 1 | 0.32% | 12/11/20 |
| R342167 | 1 | 22 | 0 | 0.00% | 22 | 0 | 0.00% | 12/11/20 |
| R325249 | 13 | 23 | 0 | 0.00% | 296 | 0 | 0.00% | 12/11/20 |
| R426748 | 1 | 9 | 0 | 0.00% | 9 | 0 | 0.00% | 12/11/20 |
| R343003 | 27 | 23 | 0 | 0.00% | 625 | 0 | 0.00% | 12/11/20 |
| R324833 | 49 | 21 | 0 | 0.00% | 1052 | 0 | 0.00% | 12/11/20 |
| R424392 | 3 | 18 | 0 | 0.00% | 55 | 0 | 0.00% | 12/11/20 |

| | | | | | | | | |
|---|---|---|---|---|---|---|---|---|
| R342986 | 3 | 19 | 0 | 0.00% | 58 | 0 | 0.00% | 12/11/20 |
| R324727 | 5 | 23 | 0 | 0.00% | 117 | 0 | 0.00% | 12/11/20 |
| R341858 | 31 | 23 | 0 | 0.00% | 727 | 0 | 0.00% | 12/11/20 |
| R339492 | 4 | 17 | 0 | 0.00% | 67 | 0 | 0.00% | 12/11/20 |
| R325529 | 6 | 23 | 0 | 0.00% | 136 | 0 | 0.00% | 12/11/20 |
| R339485 | 4 | 22 | 0 | 0.00% | 89 | 0 | 0.00% | 12/11/20 |
| R424356 | 1 | 24 | 0 | 0.00% | 24 | 0 | 0.00% | 12/11/20 |
| R326854 | 1 | 21 | 0 | 0.00% | 21 | 0 | 0.00% | 12/11/20 |
| R325125 | 4 | 20 | 2 | 50.00% | 80 | 4 | 5.00% | 12/12/20 |
| R383226 | 2 | 21 | 0 | 0.00% | 41 | 0 | 0.00% | 12/12/20 |
| R325138 | 1 | 14 | 0 | 0.00% | 14 | 0 | 0.00% | 12/12/20 |
| R424398 | 5 | 21 | 1 | 20.00% | 103 | 1 | 0.97% | 12/14/20 |
| R424410 | 10 | 22 | 1 | 10.00% | 217 | 1 | 0.46% | 12/14/20 |
| R424386 | 8 | 18 | 0 | 0.00% | 143 | 0 | 0.00% | 12/14/20 |
| R325249 | 9 | 20 | 0 | 0.00% | 176 | 0 | 0.00% | 12/14/20 |
| R325755 | 7 | 22 | 0 | 0.00% | 153 | 0 | 0.00% | 12/14/20 |
| R424428 | 1 | 17 | 0 | 0.00% | 17 | 0 | 0.00% | 12/14/20 |
| R325127 | 26 | 23 | 0 | 0.00% | 596 | 0 | 0.00% | 12/14/20 |
| R382276 | 1 | 24 | 0 | 0.00% | 24 | 0 | 0.00% | 12/14/20 |
| R325129 | 7 | 20 | 2 | 28.57% | 140 | 2 | 1.43% | 12/15/20 |
| R325305 | 54 | 23 | 2 | 3.70% | 1255 | 3 | 0.24% | 12/15/20 |
| R325529 | 6 | 21 | 1 | 16.67% | 128 | 1 | 0.78% | 12/15/20 |
| R325125 | 7 | 24 | 1 | 14.29% | 166 | 1 | 0.60% | 12/15/20 |
| R325134 | 10 | 24 | 1 | 10.00% | 238 | 1 | 0.42% | 12/15/20 |
| R339853 | 11 | 23 | 1 | 9.09% | 258 | 1 | 0.39% | 12/15/20 |
| R325127 | 14 | 23 | 1 | 7.14% | 320 | 1 | 0.31% | 12/15/20 |
| R339851 | 15 | 22 | 1 | 6.67% | 328 | 1 | 0.30% | 12/15/20 |
| R382442 | 15 | 22 | 1 | 6.67% | 336 | 1 | 0.30% | 12/15/20 |
| R325241 | 59 | 17 | 1 | 1.69% | 975 | 1 | 0.10% | 12/15/20 |
| R425408 | 64 | 24 | 1 | 1.56% | 1527 | 2 | 0.13% | 12/15/20 |
| R341849 | 13 | 23 | 0 | 0.00% | 300 | 0 | 0.00% | 12/15/20 |
| R383622 | 6 | 16 | 0 | 0.00% | 96 | 0 | 0.00% | 12/15/20 |
| R341827 | 7 | 16 | 0 | 0.00% | 111 | 0 | 0.00% | 12/15/20 |
| R339422 | 5 | 18 | 0 | 0.00% | 90 | 0 | 0.00% | 12/15/20 |
| R325139 | 9 | 23 | 0 | 0.00% | 204 | 0 | 0.00% | 12/15/20 |
| R325570 | 2 | 16 | 0 | 0.00% | 32 | 0 | 0.00% | 12/15/20 |
| R341057 | 27 | 22 | 0 | 0.00% | 590 | 0 | 0.00% | 12/15/20 |
| R339490 | 1 | 9 | 0 | 0.00% | 9 | 0 | 0.00% | 12/15/20 |
| R324836 | 56 | 15 | 4 | 7.14% | 836 | 3 | 0.36% | 12/16/20 |
| R341849 | 19 | 23 | 3 | 15.79% | 441 | 2 | 0.45% | 12/16/20 |
| R426349 | 10 | 24 | 2 | 20.00% | 237 | 2 | 0.84% | 12/16/20 |
| R339318 | 4 | 21 | 1 | 25.00% | 85 | 1 | 1.18% | 12/16/20 |
| R341850 | 13 | 20 | 1 | 7.69% | 254 | 1 | 0.39% | 12/16/20 |
| R325249 | 16 | 21 | 1 | 6.25% | 335 | 1 | 0.30% | 12/16/20 |
| R425408 | 34 | 24 | 1 | 2.94% | 812 | 1 | 0.12% | 12/16/20 |
| R341827 | 8 | 20 | 0 | 0.00% | 156 | 0 | 0.00% | 12/16/20 |
| R325554 | 14 | 20 | 0 | 0.00% | 280 | 0 | 0.00% | 12/16/20 |
| R342167 | 1 | 18 | 0 | 0.00% | 18 | 0 | 0.00% | 12/16/20 |
| R334335 | 4 | 19 | 0 | 0.00% | 74 | 0 | 0.00% | 12/16/20 |
| R341840 | 5 | 22 | 0 | 0.00% | 109 | 0 | 0.00% | 12/16/20 |
| R325755 | 9 | 19 | 0 | 0.00% | 173 | 0 | 0.00% | 12/16/20 |
| R325137 | 3 | 20 | 0 | 0.00% | 61 | 0 | 0.00% | 12/16/20 |
| R334342 | 4 | 25 | 0 | 0.00% | 98 | 0 | 0.00% | 12/16/20 |
| R342151 | 5 | 20 | 0 | 0.00% | 98 | 0 | 0.00% | 12/16/20 |
| R334324 | 3 | 18 | 0 | 0.00% | 55 | 0 | 0.00% | 12/16/20 |
| R324833 | 50 | 20 | 7 | 14.00% | 1013 | 12 | 1.18% | 12/18/20 |
| R324833 | 50 | 20 | 7 | 14.00% | 1013 | 12 | 1.18% | 12/18/20 |
| R339492 | 4 | 16 | 4 | 100.00% | 65 | 4 | 6.15% | 12/18/20 |

| | | | | | | | | |
|---|---|---|---|---|---|---|---|---|
| R339492 | 4 | 16 | 4 | 100.00% | 65 | 4 | 6.15% | 12/18/20 |
| R339483 | 8 | 18 | 2 | 25.00% | 146 | 2 | 1.37% | 12/18/20 |
| R339483 | 8 | 18 | 2 | 25.00% | 146 | 2 | 1.37% | 12/18/20 |
| R339318 | 4 | 18 | 1 | 25.00% | 71 | 2 | 2.82% | 12/19/20 |
| R339318 | 4 | 18 | 1 | 25.00% | 71 | 2 | 2.82% | 12/19/20 |
| R382276 | 1 | 24 | 1 | 100.00% | 24 | 2 | 8.33% | 12/21/20 |
| R382276 | 1 | 24 | 1 | 100.00% | 24 | 2 | 8.33% | 12/21/20 |
| R324833 | 25 | 23 | 2 | 8.00% | 563 | 4 | 0.71% | 12/23/20 |
| R324833 | 25 | 23 | 2 | 8.00% | 563 | 4 | 0.71% | 12/23/20 |
| R341866 | 28 | 24 | 7 | 25.00% | 669 | 15 | 2.24% | 1/4/21 |
| R341866 | 28 | 24 | 7 | 25.00% | 669 | 15 | 2.24% | 1/4/21 |
| R426787 | 31 | 24 | 6 | 19.35% | 739 | 12 | 1.62% | 1/4/21 |
| R426787 | 31 | 24 | 6 | 19.35% | 739 | 12 | 1.62% | 1/4/21 |
| R325554 | 4 | 21 | 3 | 75.00% | 84 | 2 | 2.38% | 1/4/21 |
| R325554 | 4 | 21 | 3 | 75.00% | 84 | 2 | 2.38% | 1/4/21 |
| R325241 | 41 | 24 | 2 | 4.88% | 972 | 10 | 1.03% | 1/4/21 |
| R325241 | 41 | 24 | 2 | 4.88% | 972 | 10 | 1.03% | 1/4/21 |
| R324833 | 7 | 24 | 1 | 14.29% | 165 | 4 | 2.42% | 1/4/21 |
| R324833 | 7 | 24 | 1 | 14.29% | 165 | 4 | 2.42% | 1/4/21 |
| R382276 | 3 | 22 | 1 | 33.33% | 66 | 6 | 9.09% | 1/4/21 |
| R382276 | 3 | 22 | 1 | 33.33% | 66 | 6 | 9.09% | 1/4/21 |
| R325125 | 92 | 24 | 10 | 10.87% | 2205 | 1 | 0.05% | 1/5/21 |
| R325125 | 92 | 24 | 10 | 10.87% | 2205 | 1 | 0.05% | 1/5/21 |
| R382442 | 32 | 23 | 3 | 9.38% | 747 | 8 | 1.07% | 1/5/21 |
| R382442 | 32 | 23 | 3 | 9.38% | 747 | 8 | 1.07% | 1/5/21 |
| R425408 | 66 | 23 | 3 | 4.55% | 1504 | 11 | 0.73% | 1/5/21 |
| R425408 | 66 | 23 | 3 | 4.55% | 1504 | 11 | 0.73% | 1/5/21 |
| R343003 | 11 | 23 | 2 | 18.18% | 250 | 4 | 1.60% | 1/5/21 |
| R343003 | 11 | 23 | 2 | 18.18% | 250 | 4 | 1.60% | 1/5/21 |
| R325129 | 9 | 21 | 2 | 22.22% | 187 | 4 | 2.14% | 1/5/21 |
| R325129 | 9 | 21 | 2 | 22.22% | 187 | 4 | 2.14% | 1/5/21 |
| R339472 | 33 | 23 | 2 | 6.06% | 772 | 9 | 1.17% | 1/5/21 |
| R339472 | 33 | 23 | 2 | 6.06% | 772 | 9 | 1.17% | 1/5/21 |
| R341827 | 55 | 19 | 1 | 1.82% | 1065 | 4 | 0.38% | 1/5/21 |
| R341827 | 55 | 19 | 1 | 1.82% | 1065 | 4 | 0.38% | 1/5/21 |
| R325148 | 6 | 20 | 1 | 16.67% | 118 | 1 | 0.85% | 1/5/21 |
| R325148 | 6 | 20 | 1 | 16.67% | 118 | 1 | 0.85% | 1/5/21 |
| R341825 | 16 | 19 | 1 | 6.25% | 306 | 4 | 1.31% | 1/5/21 |
| R341825 | 16 | 19 | 1 | 6.25% | 306 | 4 | 1.31% | 1/5/21 |
| R382253 | 29 | 24 | 10 | 34.48% | 685 | 22 | 3.21% | 1/6/21 |
| R382253 | 29 | 24 | 10 | 34.48% | 685 | 22 | 3.21% | 1/6/21 |
| R340789 | 15 | 19 | 2 | 13.33% | 292 | 4 | 1.37% | 1/6/21 |
| R340789 | 15 | 19 | 2 | 13.33% | 292 | 4 | 1.37% | 1/6/21 |
| R341850 | 13 | 20 | 2 | 15.38% | 261 | 4 | 1.53% | 1/6/21 |
| R341850 | 13 | 20 | 2 | 15.38% | 261 | 4 | 1.53% | 1/6/21 |
| R339472 | 21 | 24 | 2 | 9.52% | 501 | 3 | 0.60% | 1/6/21 |
| R339472 | 21 | 24 | 2 | 9.52% | 501 | 3 | 0.60% | 1/6/21 |
| R425408 | 34 | 23 | 2 | 5.88% | 781 | 3 | 0.38% | 1/6/21 |
| R425408 | 34 | 23 | 2 | 5.88% | 781 | 3 | 0.38% | 1/6/21 |
| R339422 | 4 | 23 | 1 | 25.00% | 92 | 4 | 4.35% | 1/6/21 |
| R339422 | 4 | 23 | 1 | 25.00% | 92 | 4 | 4.35% | 1/6/21 |
| R339308 | 2 | 19 | 1 | 50.00% | 38 | 1 | 2.63% | 1/6/21 |
| R339308 | 2 | 19 | 1 | 50.00% | 38 | 1 | 2.63% | 1/6/21 |
| R325129 | 3 | 13 | 1 | 33.33% | 39 | 2 | 5.13% | 1/6/21 |
| R325129 | 3 | 13 | 1 | 33.33% | 39 | 2 | 5.13% | 1/6/21 |
| R339318 | 4 | 20 | 1 | 25.00% | 80 | 2 | 2.50% | 1/6/21 |
| R339318 | 4 | 20 | 1 | 25.00% | 80 | 2 | 2.50% | 1/6/21 |
| R426665 | 10 | 17 | 1 | 10.00% | 172 | 2 | 1.16% | 1/6/21 |

| | | | | | | | | |
|---|---|---|---|---|---|---|---|---|
| R426665 | 10 | 17 | 1 | 10.00% | 172 | 2 | 1.16% | 1/6/21 |
| R382442 | 15 | 24 | 1 | 6.67% | 355 | 10 | 2.82% | 1/6/21 |
| R382442 | 15 | 24 | 1 | 6.67% | 355 | 10 | 2.82% | 1/6/21 |
| R325305 | 55 | 24 | 9 | 16.36% | 1323 | 27 | 2.04% | 1/7/21 |
| R325305 | 55 | 24 | 9 | 16.36% | 1323 | 27 | 2.04% | 1/7/21 |
| R423648 | 32 | 23 | 7 | 21.88% | 739 | 13 | 1.76% | 1/7/21 |
| R423648 | 32 | 23 | 7 | 21.88% | 739 | 13 | 1.76% | 1/7/21 |
| R382442 | 19 | 24 | 7 | 36.84% | 448 | 21 | 4.69% | 1/7/21 |
| R382442 | 19 | 24 | 7 | 36.84% | 448 | 21 | 4.69% | 1/7/21 |
| R324836 | 59 | 17 | 4 | 6.78% | 1007 | 14 | 1.39% | 1/7/21 |
| R324836 | 59 | 17 | 4 | 6.78% | 1007 | 14 | 1.39% | 1/7/21 |
| R325134 | 14 | 23 | 1 | 7.14% | 327 | 2 | 0.61% | 1/7/21 |
| R325134 | 14 | 23 | 1 | 7.14% | 327 | 2 | 0.61% | 1/7/21 |
| R325249 | 5 | 19 | 1 | 20.00% | 97 | 2 | 2.06% | 1/7/21 |
| R325249 | 5 | 19 | 1 | 20.00% | 97 | 2 | 2.06% | 1/7/21 |
| R334335 | 4 | 20 | 1 | 25.00% | 80 | 2 | 2.50% | 1/7/21 |
| R334335 | 4 | 20 | 1 | 25.00% | 80 | 2 | 2.50% | 1/7/21 |
| R426349 | 11 | 23 | 1 | 9.09% | 253 | 4 | 1.58% | 1/7/21 |
| R426349 | 11 | 23 | 1 | 9.09% | 253 | 4 | 1.58% | 1/7/21 |
| R324833 | 50 | 21 | 5 | 10.00% | 1062 | 12 | 1.13% | 1/8/21 |
| R324833 | 50 | 21 | 5 | 10.00% | 1062 | 12 | 1.13% | 1/8/21 |
| R324851 | 63 | 24 | 3 | 4.76% | 1506 | 6 | 0.40% | 1/8/21 |
| R324851 | 63 | 24 | 3 | 4.76% | 1506 | 6 | 0.40% | 1/8/21 |
| R325125 | 38 | 24 | 3 | 7.89% | 901 | 1 | 0.11% | 1/8/21 |
| R325125 | 38 | 24 | 3 | 7.89% | 901 | 1 | 0.11% | 1/8/21 |
| R326820 | 13 | 22 | 2 | 15.38% | 290 | 3 | 1.03% | 1/8/21 |
| R326820 | 13 | 22 | 2 | 15.38% | 290 | 3 | 1.03% | 1/8/21 |
| R424410 | 10 | 21 | 1 | 10.00% | 208 | 1 | 0.48% | 1/8/21 |
| R424410 | 10 | 21 | 1 | 10.00% | 208 | 1 | 0.48% | 1/8/21 |
| R325127 | 3 | 23 | 1 | 33.33% | 69 | 2 | 2.90% | 1/8/21 |
| R325127 | 3 | 23 | 1 | 33.33% | 69 | 2 | 2.90% | 1/8/21 |
| R339483 | 8 | 19 | 1 | 12.50% | 152 | 2 | 1.32% | 1/8/21 |
| R339483 | 8 | 19 | 1 | 12.50% | 152 | 2 | 1.32% | 1/8/21 |
| R334342 | 13 | 23 | 1 | 7.69% | 305 | 2 | 0.66% | 1/8/21 |
| R334342 | 13 | 23 | 1 | 7.69% | 305 | 2 | 0.66% | 1/8/21 |
| R325529 | 4 | 13 | 2 | 50.00% | 53 | 4 | 7.55% | 1/9/21 |
| R325529 | 4 | 13 | 2 | 50.00% | 53 | 4 | 7.55% | 1/9/21 |
| R324833 | 1 | 24 | 1 | 100.00% | 24 | 2 | 8.33% | 1/9/21 |
| R324833 | 1 | 24 | 1 | 100.00% | 24 | 2 | 8.33% | 1/9/21 |
| R334342 | 5 | 23 | 1 | 20.00% | 113 | 1 | 0.88% | 1/9/21 |
| R334342 | 5 | 23 | 1 | 20.00% | 113 | 1 | 0.88% | 1/9/21 |
| R325755 | 8 | 21 | 1 | 12.50% | 169 | 1 | 0.59% | 1/11/21 |
| R325755 | 8 | 21 | 1 | 12.50% | 169 | 1 | 0.59% | 1/11/21 |
| R425408 | 62 | 24 | 2 | 3.23% | 1458 | 2 | 0.14% | 1/12/21 |
| R425408 | 62 | 24 | 2 | 3.23% | 1458 | 2 | 0.14% | 1/12/21 |
| R325136 | 16 | 23 | 1 | 6.25% | 361 | 1 | 0.28% | 1/12/21 |
| R325136 | 16 | 23 | 1 | 6.25% | 361 | 1 | 0.28% | 1/12/21 |
| R325249 | 14 | 21 | 2 | 14.29% | 298 | 3 | 1.01% | 1/13/21 |
| R325249 | 14 | 21 | 2 | 14.29% | 298 | 3 | 1.01% | 1/13/21 |
