## Supplementary Table 3 for "Pooled Surveillance Testing Program for Asymptomatic SARS-CoV-2 Infections in K-12 Schools and Universities"

Supplementary Table 3. Raw data of each university tested between Sept 1, 2020 and Jan 13, 2021.

| University NO. | Pool number | Pool size | Pool Positive number | Pool Positivity rate | Individual number | Individual Positive number | Individual Positivity rate | Date |
|---|---|---|---|---|---|---|---|---|
| R326848 | 10 | 24 | 0 | 0.00% | 240 | 0 | 0.00% | 9/1/20 |
| R326848 | 12 | 23 | 0 | 0.00% | 277 | 0 | 0.00% | 9/2/20 |
| R326848 | 17 | 21 | 0 | 0.00% | 365 | 0 | 0.00% | 9/3/20 |
| R326848 | 11 | 18 | 0 | 0.00% | 199 | 0 | 0.00% | 9/10/20 |
| R326848 | 2 | 24 | 0 | 0.00% | 48 | 0 | 0.00% | 9/15/20 |
| R326848 | 8 | 22 | 4 | 50.00% | 179 | 2 | 1.12% | 9/16/20 |
| R326848 | 5 | 24 | 0 | 0.00% | 121 | 0 | 0.00% | 9/22/20 |
| R325293 | 10 | 23 | 5 | 50.00% | 225 | 8 | 3.56% | 9/24/20 |
| R325302 | 3 | 20 | 0 | 0.00% | 61 | 0 | 0.00% | 9/26/20 |
| R326848 | 6 | 22 | 0 | 0.00% | 134 | 0 | 0.00% | 9/29/20 |
| R325302 | 1 | 18 | 0 | 0.00% | 18 | 0 | 0.00% | 9/29/20 |
| R325293 | 5 | 25 | 1 | 20.00% | 124 | 1 | 0.81% | 9/30/20 |
| R325283 | 1 | 24 | 0 | 0.00% | 24 | 0 | 0.00% | 9/30/20 |
| R325269 | 6 | 22 | 0 | 0.00% | 131 | 0 | 0.00% | 9/30/20 |
| R325131 | 10 | 22 | 1 | 10.00% | 223 | 1 | 0.45% | 10/1/20 |
| R325293 | 2 | 25 | 0 | 0.00% | 50 | 0 | 0.00% | 10/1/20 |
| R325293 | 4 | 24 | 0 | 0.00% | 96 | 0 | 0.00% | 10/2/20 |
| R325302 | 5 | 22 | 3 | 60.00% | 110 | 2 | 1.82% | 10/3/20 |
| R325130 | 4 | 24 | 0 | 0.00% | 94 | 0 | 0.00% | 10/6/20 |
| R326848 | 9 | 21 | 0 | 0.00% | 189 | 0 | 0.00% | 10/6/20 |
| R325283 | 1 | 24 | 0 | 0.00% | 24 | 0 | 0.00% | 10/7/20 |
| R325170 | 1 | 23 | 0 | 0.00% | 23 | 0 | 0.00% | 10/7/20 |
| R325302 | 1 | 9 | 0 | 0.00% | 9 | 0 | 0.00% | 10/8/20 |
| R325269 | 2 | 22 | 0 | 0.00% | 43 | 0 | 0.00% | 10/8/20 |
| R326848 | 2 | 20 | 0 | 0.00% | 39 | 0 | 0.00% | 10/9/20 |
| R325130 | 2 | 23 | 0 | 0.00% | 46 | 0 | 0.00% | 10/13/20 |
| R326848 | 4 | 22 | 0 | 0.00% | 87 | 0 | 0.00% | 10/13/20 |
| R325283 | 1 | 24 | 0 | 0.00% | 24 | 0 | 0.00% | 10/14/20 |
| R325269 | 2 | 23 | 0 | 0.00% | 46 | 0 | 0.00% | 10/14/20 |
| R325302 | 2 | 17 | 0 | 0.00% | 33 | 0 | 0.00% | 10/15/20 |
| R325131 | 5 | 23 | 0 | 0.00% | 115 | 0 | 0.00% | 10/15/20 |
| R325293 | 13 | 23 | 0 | 0.00% | 305 | 0 | 0.00% | 10/19/20 |
| R325130 | 2 | 24 | 0 | 0.00% | 48 | 0 | 0.00% | 10/20/20 |
| R325536 | 6 | 23 | 0 | 0.00% | 136 | 0 | 0.00% | 10/20/20 |
| R325302 | 1 | 5 | 0 | 0.00% | 5 | 0 | 0.00% | 10/20/20 |
| R325293 | 6 | 19 | 0 | 0.00% | 112 | 0 | 0.00% | 10/20/20 |
| R325283 | 1 | 24 | 0 | 0.00% | 24 | 0 | 0.00% | 10/21/20 |
| R326848 | 8 | 24 | 0 | 0.00% | 192 | 0 | 0.00% | 10/21/20 |

| | | | | | | | | |
|---|---|---|---|---|---|---|---|---|
| R425660 | 10 | 21 | 0 | 0.00% | 210 | 0 | 0.00% | 10/22/20 |
| R426905 | 1 | 24 | 0 | 0.00% | 24 | 0 | 0.00% | 10/23/20 |
| R326848 | 6 | 22 | 0 | 0.00% | 130 | 0 | 0.00% | 10/27/20 |
| R325536 | 7 | 23 | 0 | 0.00% | 162 | 0 | 0.00% | 10/27/20 |
| R325302 | 1 | 15 | 0 | 0.00% | 15 | 0 | 0.00% | 10/27/20 |
| R325170 | 4 | 22 | 0 | 0.00% | 87 | 0 | 0.00% | 10/27/20 |
| R325283 | 1 | 24 | 0 | 0.00% | 24 | 0 | 0.00% | 10/28/20 |
| R325130 | 4 | 23 | 0 | 0.00% | 90 | 0 | 0.00% | 10/28/20 |
| R325269 | 2 | 24 | 0 | 0.00% | 48 | 0 | 0.00% | 10/28/20 |
| R325131 | 5 | 21 | 0 | 0.00% | 103 | 0 | 0.00% | 10/29/20 |
| R325302 | 1 | 24 | 0 | 0.00% | 24 | 0 | 0.00% | 10/30/20 |
| R325170 | 2 | 22 | 0 | 0.00% | 43 | 0 | 0.00% | 11/2/20 |
| R325536 | 9 | 23 | 1 | 11.11% | 204 | 1 | 0.49% | 11/3/20 |
| R325130 | 4 | 24 | 0 | 0.00% | 94 | 0 | 0.00% | 11/3/20 |
| R326848 | 9 | 21 | 0 | 0.00% | 191 | 0 | 0.00% | 11/3/20 |
| R325302 | 2 | 20 | 0 | 0.00% | 40 | 0 | 0.00% | 11/3/20 |
| R325170 | 8 | 21 | 2 | 25.00% | 166 | 4 | 2.41% | 11/4/20 |
| R325283 | 1 | 24 | 0 | 0.00% | 24 | 0 | 0.00% | 11/4/20 |
| R325302 | 2 | 18 | 0 | 0.00% | 36 | 0 | 0.00% | 11/5/20 |
| R325293 | 2 | 25 | 0 | 0.00% | 49 | 0 | 0.00% | 11/5/20 |
| R326848 | 2 | 21 | 0 | 0.00% | 42 | 0 | 0.00% | 11/6/20 |
| R325302 | 2 | 17 | 0 | 0.00% | 34 | 0 | 0.00% | 11/7/20 |
| R325170 | 4 | 17 | 0 | 0.00% | 67 | 0 | 0.00% | 11/9/20 |
| R325130 | 2 | 24 | 0 | 0.00% | 47 | 0 | 0.00% | 11/10/20 |
| R326848 | 7 | 23 | 0 | 0.00% | 162 | 0 | 0.00% | 11/10/20 |
| R325536 | 11 | 22 | 0 | 0.00% | 244 | 0 | 0.00% | 11/10/20 |
| R325302 | 2 | 21 | 0 | 0.00% | 41 | 0 | 0.00% | 11/10/20 |
| R325293 | 2 | 24 | 0 | 0.00% | 47 | 0 | 0.00% | 11/10/20 |
| R325170 | 33 | 24 | 14 | 42.42% | 781 | 16 | 2.05% | 11/11/20 |
| R325269 | 2 | 21 | 2 | 100.00% | 41 | 2 | 4.88% | 11/11/20 |
| R325283 | 1 | 24 | 0 | 0.00% | 24 | 0 | 0.00% | 11/11/20 |
| R325131 | 5 | 24 | 0 | 0.00% | 120 | 0 | 0.00% | 11/12/20 |
| R341828 | 6 | 22 | 4 | 66.67% | 132 | 26 | 19.70% | 11/13/20 |
| R325302 | 2 | 19 | 0 | 0.00% | 38 | 0 | 0.00% | 11/13/20 |
| R425660 | 5 | 18 | 1 | 20.00% | 89 | 5 | 5.62% | 11/16/20 |
| R325302 | 2 | 17 | 0 | 0.00% | 34 | 0 | 0.00% | 11/16/20 |
| R340810 | 2 | 18 | 0 | 0.00% | 36 | 0 | 0.00% | 11/16/20 |
| R341869 | 2 | 20 | 0 | 0.00% | 39 | 0 | 0.00% | 11/16/20 |
| R325536 | 24 | 16 | 2 | 8.33% | 395 | 4 | 1.01% | 11/17/20 |
| R425660 | 12 | 21 | 1 | 8.33% | 249 | 1 | 0.40% | 11/17/20 |
| R326848 | 10 | 21 | 0 | 0.00% | 210 | 0 | 0.00% | 11/17/20 |
| R325302 | 2 | 20 | 0 | 0.00% | 39 | 0 | 0.00% | 11/17/20 |
| R340810 | 2 | 19 | 0 | 0.00% | 37 | 0 | 0.00% | 11/17/20 |

| | | | | | | | | |
|---|---|---|---|---|---|---|---|---|
| R325293 | 12 | 24 | 0 | 0.00% | 285 | 0 | 0.00% | 11/17/20 |
| R325130 | 4 | 23 | 1 | 25.00% | 92 | 1 | 1.09% | 11/18/20 |
| R325283 | 2 | 21 | 0 | 0.00% | 41 | 0 | 0.00% | 11/18/20 |
| R341828 | 5 | 24 | 5 | 100.00% | 120 | 6 | 5.00% | 11/19/20 |
| R325302 | 2 | 19 | 1 | 50.00% | 37 | 2 | 5.41% | 11/19/20 |
| R325131 | 7 | 22 | 0 | 0.00% | 152 | 0 | 0.00% | 11/19/20 |
| R341869 | 2 | 12 | 0 | 0.00% | 24 | 0 | 0.00% | 11/19/20 |
| R325536 | 7 | 17 | 0 | 0.00% | 120 | 0 | 0.00% | 11/20/20 |
| R325302 | 1 | 25 | 0 | 0.00% | 25 | 0 | 0.00% | 11/23/20 |
| R340810 | 2 | 19 | 0 | 0.00% | 37 | 0 | 0.00% | 11/23/20 |
| R341869 | 2 | 19 | 0 | 0.00% | 37 | 0 | 0.00% | 11/23/20 |
| R325269 | 2 | 20 | 2 | 100.00% | 39 | 2 | 5.13% | 11/24/20 |
| R326848 | 9 | 23 | 0 | 0.00% | 203 | 0 | 0.00% | 11/24/20 |
| R325302 | 2 | 14 | 0 | 0.00% | 28 | 0 | 0.00% | 11/24/20 |
| R339475 | 1 | 24 | 0 | 0.00% | 24 | 0 | 0.00% | 11/24/20 |
| R341828 | 1 | 23 | 0 | 0.00% | 23 | 0 | 0.00% | 11/24/20 |
| R340810 | 2 | 18 | 0 | 0.00% | 35 | 0 | 0.00% | 11/25/20 |
| R341869 | 2 | 19 | 0 | 0.00% | 38 | 0 | 0.00% | 11/30/20 |
| R325283 | 2 | 21 | 1 | 50.00% | 41 | 1 | 2.44% | 12/1/20 |
| R325130 | 2 | 23 | 0 | 0.00% | 45 | 0 | 0.00% | 12/1/20 |
| R325536 | 15 | 24 | 0 | 0.00% | 353 | 0 | 0.00% | 12/1/20 |
| R341830 | 2 | 18 | 0 | 0.00% | 35 | 0 | 0.00% | 12/1/20 |
| R325302 | 2 | 20 | 0 | 0.00% | 39 | 0 | 0.00% | 12/1/20 |
| R340810 | 3 | 16 | 0 | 0.00% | 47 | 0 | 0.00% | 12/1/20 |
| R341830 | 2 | 18 | 0 | 0.00% | 36 | 0 | 0.00% | 12/2/20 |
| R325131 | 3 | 21 | 0 | 0.00% | 64 | 0 | 0.00% | 12/2/20 |
| R325170 | 6 | 22 | 0 | 0.00% | 134 | 0 | 0.00% | 12/2/20 |
| R340810 | 2 | 26 | 0 | 0.00% | 52 | 0 | 0.00% | 12/2/20 |
| R423642 | 1 | 20 | 0 | 0.00% | 20 | 0 | 0.00% | 12/3/20 |
| R326848 | 13 | 22 | 0 | 0.00% | 282 | 0 | 0.00% | 12/3/20 |
| R325302 | 2 | 15 | 0 | 0.00% | 30 | 0 | 0.00% | 12/3/20 |
| R326848 | 6 | 22 | 0 | 0.00% | 134 | 0 | 0.00% | 12/4/20 |
| R341830 | 2 | 14 | 0 | 0.00% | 27 | 0 | 0.00% | 12/4/20 |
| R325131 | 2 | 20 | 0 | 0.00% | 39 | 0 | 0.00% | 12/4/20 |
| R340810 | 2 | 16 | 0 | 0.00% | 32 | 0 | 0.00% | 12/4/20 |
| R341828 | 2 | 24 | 0 | 0.00% | 47 | 0 | 0.00% | 12/4/20 |
| R341830 | 2 | 18 | 0 | 0.00% | 36 | 0 | 0.00% | 12/5/20 |
| R325170 | 1 | 22 | 0 | 0.00% | 22 | 0 | 0.00% | 12/5/20 |
| R340810 | 1 | 13 | 0 | 0.00% | 13 | 0 | 0.00% | 12/5/20 |
| R424416 | 8 | 24 | 3 | 37.50% | 192 | 3 | 1.56% | 12/7/20 |
| R325302 | 2 | 17 | 0 | 0.00% | 33 | 0 | 0.00% | 12/7/20 |
| R325131 | 2 | 20 | 0 | 0.00% | 40 | 0 | 0.00% | 12/7/20 |
| R341869 | 2 | 20 | 0 | 0.00% | 39 | 0 | 0.00% | 12/7/20 |

| | | | | | | | | |
|---|---|---|---|---|---|---|---|---|
| R339475 | 3 | 24 | 1 | 33.33% | 72 | 1 | 1.39% | 12/8/20 |
| R325283 | 1 | 10 | 0 | 0.00% | 10 | 0 | 0.00% | 12/8/20 |
| R325130 | 1 | 21 | 0 | 0.00% | 21 | 0 | 0.00% | 12/8/20 |
| R326848 | 10 | 23 | 0 | 0.00% | 227 | 0 | 0.00% | 12/8/20 |
| R325536 | 4 | 22 | 0 | 0.00% | 88 | 0 | 0.00% | 12/8/20 |
| R341830 | 2 | 18 | 0 | 0.00% | 36 | 0 | 0.00% | 12/8/20 |
| R325302 | 2 | 24 | 0 | 0.00% | 48 | 0 | 0.00% | 12/8/20 |
| R340810 | 2 | 19 | 0 | 0.00% | 37 | 0 | 0.00% | 12/8/20 |
| R339475 | 3 | 24 | 0 | 0.00% | 72 | 0 | 0.00% | 12/9/20 |
| R339475 | 3 | 24 | 1 | 33.33% | 72 | 1 | 1.39% | 12/10/20 |
| R341830 | 2 | 18 | 0 | 0.00% | 36 | 0 | 0.00% | 12/10/20 |
| R325302 | 2 | 19 | 0 | 0.00% | 37 | 0 | 0.00% | 12/10/20 |
| R325131 | 2 | 20 | 0 | 0.00% | 40 | 0 | 0.00% | 12/10/20 |
| R340810 | 3 | 19 | 0 | 0.00% | 58 | 0 | 0.00% | 12/10/20 |
| R423642 | 1 | 21 | 0 | 0.00% | 21 | 0 | 0.00% | 12/11/20 |
| R325131 | 2 | 20 | 0 | 0.00% | 40 | 0 | 0.00% | 12/11/20 |
| R339475 | 3 | 24 | 0 | 0.00% | 72 | 0 | 0.00% | 12/11/20 |
| R341830 | 1 | 17 | 0 | 0.00% | 17 | 0 | 0.00% | 12/12/20 |
| R339475 | 3 | 25 | 0 | 0.00% | 76 | 0 | 0.00% | 12/12/20 |
| R326848 | 5 | 24 | 1 | 20.00% | 119 | 1 | 0.84% | 12/15/20 |
| R325130 | 1 | 21 | 0 | 0.00% | 21 | 0 | 0.00% | 12/15/20 |
| R341830 | 1 | 13 | 0 | 0.00% | 13 | 0 | 0.00% | 12/15/20 |
| R325302 | 2 | 24 | 0 | 0.00% | 47 | 0 | 0.00% | 12/15/20 |
| R340810 | 2 | 19 | 0 | 0.00% | 38 | 0 | 0.00% | 12/15/20 |
| R325170 | 6 | 18 | 0 | 0.00% | 106 | 0 | 0.00% | 12/16/20 |
| R423642 | 1 | 24 | 1 | 100.00% | 24 | 17 | 70.83% | 12/17/20 |
| R423642 | 1 | 24 | 1 | 100.00% | 24 | 17 | 70.83% | 12/17/20 |
| R423642 | 1 | 24 | 1 | 100.00% | 24 | 17 | 70.83% | 12/17/20 |
| R423642 | 1 | 24 | 1 | 100.00% | 24 | 17 | 70.83% | 12/17/20 |
| R339475 | 5 | 23 | 2 | 40.00% | 114 | 8 | 7.02% | 12/19/20 |
| R339475 | 5 | 23 | 2 | 40.00% | 114 | 8 | 7.02% | 12/19/20 |
| R325536 | 13 | 23 | 1 | 7.69% | 300 | 4 | 1.33% | 1/5/21 |
| R325536 | 13 | 23 | 1 | 7.69% | 300 | 4 | 1.33% | 1/5/21 |
| R325302 | 4 | 23 | 1 | 25.00% | 92 | 4 | 4.35% | 1/5/21 |
| R325302 | 4 | 23 | 1 | 25.00% | 92 | 4 | 4.35% | 1/5/21 |
| R340810 | 1 | 24 | 1 | 100.00% | 24 | 4 | 16.67% | 1/5/21 |
| R340810 | 1 | 24 | 1 | 100.00% | 24 | 4 | 16.67% | 1/5/21 |
| R325131 | 4 | 19 | 1 | 25.00% | 76 | 2 | 2.63% | 1/6/21 |
| R325131 | 4 | 19 | 1 | 25.00% | 76 | 2 | 2.63% | 1/6/21 |
| R325130 | 12 | 21 | 4 | 33.33% | 254 | 18 | 7.09% | 1/7/21 |
| R325130 | 12 | 21 | 4 | 33.33% | 254 | 18 | 7.09% | 1/7/21 |
| R325302 | 6 | 17 | 3 | 50.00% | 104 | 10 | 9.62% | 1/7/21 |
| R325302 | 6 | 17 | 3 | 50.00% | 104 | 10 | 9.62% | 1/7/21 |

| | | | | | | | | |
|---|---|---|---|---|---|---|---|---|
| R326848 | 7 | 23 | 1 | 14.29% | 159 | 2 | 1.26% | 1/7/21 |
| R326848 | 7 | 23 | 1 | 14.29% | 159 | 2 | 1.26% | 1/7/21 |
| R325131 | 6 | 21 | 2 | 33.33% | 125 | 8 | 6.40% | 1/8/21 |
| R325131 | 6 | 21 | 2 | 33.33% | 125 | 8 | 6.40% | 1/8/21 |
| R325130 | 3 | 21 | 1 | 33.33% | 64 | 2 | 3.13% | 1/8/21 |
| R325130 | 3 | 21 | 1 | 33.33% | 64 | 2 | 3.13% | 1/8/21 |
| R325536 | 36 | 24 | 8 | 22.22% | 853 | 11 | 1.29% | 1/12/21 |
| R325536 | 36 | 24 | 8 | 22.22% | 853 | 11 | 1.29% | 1/12/21 |
| R325130 | 8 | 23 | 2 | 25.00% | 185 | 3 | 1.62% | 1/12/21 |
| R325130 | 8 | 23 | 2 | 25.00% | 185 | 3 | 1.62% | 1/12/21 |
| R325302 | 5 | 24 | 1 | 20.00% | 119 | 1 | 0.84% | 1/12/21 |
| R325302 | 5 | 24 | 1 | 20.00% | 119 | 1 | 0.84% | 1/12/21 |
| R325131 | 6 | 24 | 3 | 50.00% | 142 | 4 | 2.82% | 1/13/21 |
| R325131 | 6 | 24 | 3 | 50.00% | 142 | 4 | 2.82% | 1/13/21 |
| R339475 | 4 | 19 | 1 | 25.00% | 76 | 1 | 1.32% | 1/13/21 |
| R339475 | 4 | 19 | 1 | 25.00% | 76 | 1 | 1.32% | 1/13/21 |
