## Supplementary Table 4Supplementary Table for "Pooled Surveillance Testing Program for Asymptomatic SARS-CoV-2 Infections in K-12 Schools and Universities"

Supplementary Table 4. Representative comparison of Ct values of individual specimens and pools of 24. Pools of 24 samples were generated and tested for SARS-CoV-2 viral RNA. Pools with a single gene amplification were triggered for reflex to identify the individual positive samples(s) within the pool. Pools of 24 with a single positive sample identified were selected.

| | Pool Mean CT values (SD); N=3 | | | | | Individual Mean CT values (SD); N=3 | | | |
|---|---|---|---|---|---|---|---|---|---|
| **Pool NO.** | **MS2 Phage** | **N gene** | **Orf1ab gene** | **S gene** | **Individual NO.** | **MS2 Phage** | **N gene** | **Orf1ab gene** | **S gene** |
| 02426781 | 26.83 (0.77) | 29.6 1(2.08) | 31.62 (0.65) | 30.07 (1.40) | 00209967 | 24.86 (0.12) | 27.14 (0.20) | 27.34 (0.24) | 26.94 (0.38) |
| 08146338 | 26.91 (0.99) | 30.11 (0.79) | 31.88 (1.26) | 30.25 (2.39) | 00056783 | 24.91 (0.72) | 27.26 (0.30) | 28.22 (0.11) | 28.45 (0.52) |
| 02343926 | 25.87 (0.41) | 29.97(0.03) | 29.26 (0.11) | 29.90 (0.09) | 02537656 | 28.72 (0.10) | 27.58 (0.08) | 28.17 (0.13) | 27.43 (0.08) |
| 02424449 | 27.38 (0.84) | 30.24 (0.08) | 30.57 (0.63) | 30.43 (0.60) | 00033197 | 25.04 (0.09) | 27.61 (0.16) | 26.99 (0.40) | 26.58 (0.19) |
| 02425412 | 26.45 (0.27) | 30.87 (0.84) | 33.60 (0.39) | 32.31 (0.22) | 00936516 | 24.72 (1.15) | 28.58 (1.05) | 27.08 (2.24) | 21.71 (6.65) |
| 02342504 | 25.48 (0.44) | 31.59 (NA) | 34.39 (0.92) | 34.08 (NA) | 02572290 | 24.34 (1.08) | 28.58 (1.66) | 28.82 (1.97) | 25.94 (5.27) |
| 08144478 | 29.55 (0.07) | 32.42 (0.62) | 33.15 (0.58) | 32.49 (0.68) | 00072563 | 25.44 (0.18) | 28.87 (0.12) | 29.26 (0.01) | 29.80 (0.24) |
| 02339381 | 27.11 (0.18) | 30.42 (0.46) | 31.39 (0.44) | 31.00 (0.53) | 02542540 | 27.10 (0.75) | 28.94 (0.24) | 28.25 (0.36) | 27.96 (0.37) |
| 02342514 | 25.88 (0.32) | 30.61 (NA) | 34.25 (0.30) | 31.38 (0.22) | 02570484 | 25.37 (0.66) | 29.06 (0.32) | 29.96 (0.35) | 29.08 (0.44) |
| 02343358 | 26.09 (0.53) | 34.78 (NA) | 33.43 (2.40) | 38.20 (NA) | 02601428 | 26.40 (0.26) | 29.08 (0.61) | 32.40 (0.41) | 31.86 (0.50) |
| 07211899 | 27.50 (1.88) | 30.71 (0.13) | 32.52 (0.16) | 31.66 (1.62) | 07211857 | 29.48 (0.85) | 29.42 (0.37) | 30.95 (0.33) | 29.75 (0.50) |
| 02423430 | 23.60 (0.64) | 32.44 (1.18) | 31.91 (0.94) | 31.82 (0.82) | 00314486 | 25.46 (0.22) | 29.62 (0.03) | 28.98 (0.18) | 28.46 (0.20) |
| 02339514 | 26.68 (1.24) | 31.56 (NA) | 36.01 (NA) | 31.14 (NA) | 02621847 | 26.19 (1.05) | 29.79 (0.68) | 30.57 (1.00) | 28.09 (3.16) |
| 02422747 | 31.42 (0.50) | 34.08 (NA) | 33.00 (0.35) | 33.97 (1.18) | 02571625 | 25.87 (0.36) | 29.91 (0.05) | 31.13 (0.08) | 30.45 (0.06) |
| 02382707 | 28.36 (0.52) | 33.43 (0.04) | 35.66 (0.92) | 35.20 (NA) | 00240737 | 24.50 (0.06) | 29.93 (0.61) | 30.58 (0.35) | 29.73 (0.53) |
| 02343089 | 30.79 (0.15) | 33.01 (0.07) | 33.42 (0.53) | 34.65 (1.20) | 00968143 | 27.38 (0.41) | 30.22 (0.19) | 29.69 (0.28) | 30.56 (0.45) |
| 02340249 | 28.81 (0.27) | 32.09 (1.14) | 34.96 (1.12) | 34.78 (0.21) | 00913706 | 25.84 (0.2) | 31.01 (0.37) | 30.97 (0.05) | 31.23 (0.34) |
| 02341926 | 27.67 (0.35) | 34.25 (NA) | 34.46 (NA) | 34.88 (0.31) | 00039323 | 24.84 (2.95) | 31.09 (0.27) | 30.19 (0.66) | 28.43 (1.98) |
| 02343186 | 28.65 (0.42) | 33.06 (NA) | 34.5 (NA) | 35.59 (0.54) | 02585218 | 25.84 (0.6) | 31.16 (1.02) | 31.97 (0.92) | 32.26 (NA) |
| 02340362 | 27.30 (1.12) | 34.07 (0.20) | 34.94 (NA) | 34.04 (NA) | 02583435 | 26.58 (0.21) | 31.52 (0.34) | 32.17 (0.25) | 32.14 (1.09) |
