## Supplementary Table 5 for "Pooled Surveillance Testing Program for Asymptomatic SARS-CoV-2 Infections in K-12 Schools and Universities"

Supplementary Table 5. CT values of samples stored at room temperature or cycled through worst case shipping conditions using winter or summer excursion temperatures. A winter excursion consisted of cycling from -10°C for 8 hours, 18°C for 4 hours, -10°C for 2 hours, 10°C for 36 hours, and -10°C for 6 hours before testing. A summer excursion cycled from 40°C for 8 hours, 22°C for 4 hours, 40°C for 2 hours, 30°C for 36 hours, and 40°C for 6 hours before testing.

| **Conditions** | **N** | **Mean CT values (SD; N=3)** | | | |
|---|---|---|---|---|---|
| | | **MS2 Phage** | **N gene** | **ORF1ab** | **S gene** |
| Fresh | 3 | 25.73 (0.44) | 27.37 (0.24) | 28.8 (0.23) | 27.82 (0.3) |
| RT 5 days | 10 | 26.47 (0.03) | 29.03 (0.19) | 28.27 (0.21) | 30.02 (0.24) |
| RT 10 days | 10 | 25.7 (0.14) | 26.32 (0.15) | 24.53 (0.52) | 26.52 (0.10) |
| Winter | 20 | 29.48 (1.79) | 28.41 (0.46) | 27.09 (1.03) | 25.93 (0.94) |
| Summer | 12 | 28.61 (1.07) | 28.79 (0.57) | 27.83 (0.85) | 28.08 (1.23) |
| Fresh | 3 | 25.62 (0.55) | 26.87 (0.72) | 27.2 (0.32) | 27.58 (0.34) |
| RT 5 days | 10 | 26.12 (0.14) | 26.33 (0.07) | 25.46 (0.01) | 27.34 (0.08) |
| RT 10 days | 10 | 25.14 (0.45) | 28.30 (0.19) | 27.04 (0.16) | 28.76 (0.14) |
| Winter | 15 | 28.25 (0.8) | 28.52 (0.61) | 27.57 (0.84) | 28.01 (0.9) |
| Summer | 8 | 28.02 (0.51) | 28.76 (0.48) | 27.66 (0.72) | 26.01 (2.27) |
